# Targeting Molecular Collagen Defects from the Initiation of Knee Osteoarthritis

**DOI:** 10.1101/2024.06.13.24308739

**Authors:** Kui Huang, Rongmao Qiu, Yijie Fang, Dantian Zhu, Xiaojing Li, Zhongyang Lv, Taoyu Jia, Yuxiang Fei, Daoning Zhou, Wenjun Wu, Yongjie Huang, Suwen Zhao, Yongqiao Zhu, Shaolin Li, Dongquan Shi, Yang Li

## Abstract

Knee osteoarthritis (OA) is the most prevalent degenerative joint disease. When morphological changes become apparent on radiographs, no approved treatment can reverse the disease process. Early diagnosis is an unmet need demanding new molecular and imaging biomarkers to define OA from the earliest stages. In this context, we focus on collagen, the most basic building block of all joint tissues, and interrogate how OA development affects collagen’s molecular folding, a previously underexplored area. Here, through whole-joint mapping with a peptide that recognizes unfolded collagen molecules, we report the discovery of collagen denaturation in cartilage before proteolysis and major histopathological degeneration in animal models and patients. Mechanistically, we reveal that such molecular collagen defects can be driven by mechanical overloading without collagenase degradation and are intimately associated with glycosaminoglycan loss. We showcase the advantages of using collagen denaturation as an early-stage OA hallmark for in vivo therapeutic evaluation and molecular magnetic resonance imaging (MRI) of subtle joint defects that are challenging to detect with conventional morphology-based MRI. These results highlight biomolecular integrity as a crucial dimension for characterizing joint degeneration and a molecular foundation for diagnosing early-stage OA and beyond.

## INTRODUCTION

Osteoarthritis (OA) is the most prevalent joint disease affecting over 500 million individuals worldwide, including over 365 million with knee OA.^1^ ^2^ OA is a leading cause of disability in elders and its incidence is increasing globally with aging populations and rising rates of obesity and injury.^1–3^ There is no approved disease-modifying treatment for OA.^4^ Standard radiographic changes are indirect and insensitive,^5^ such as joint-space narrowing, which may not appear for years after the disease onset.^6^ Routine OA management often focuses on progressing and late-stage patients, when joint replacement is often the ultimate recommendation.^7^ Nonetheless, OA is a slowly progressing disease; a ‘window of opportunity’ for early diagnosis and treatment does exist but remains to be precisely defined.^7^ ^8^ Therefore, new molecular and imaging biomarkers that enable accurate characterization and non-invasive examination of early-stage OA are critically needed to support a paradigm shift from reaction to late disease toward prevention.^8^

The destruction of articular cartilage is the defining characteristic of OA.^9^ The rich collagen network primarily maintains the structural integrity and mechanical functioning of cartilage and other joint tissues, including the menisci and ligaments.^10^ As OA advances, mechanical insults and increased matrix-protease activities induced by inflammatory cytokines and abnormal cell metabolism disrupt the collagen matrix progressively, leading to structural failure of the tissues and painful disorders.^1^ ^9^ ^11^ The collagen molecule is the basic building block of the hierarchical collagen architecture in cartilage.^10^ We hypothesized that the structural disruption to the collagen matrix occurs at and progresses from the molecular level in OA from the initiation.

The molecular defects in collagen structure, namely the denaturation and unwinding of its unique triple-helix folding, cannot be directly detected by regular imaging, histology, and electron microscopies.^12^ Alternatively, current research for collagen changes in OA focuses on matrix proteases,^13^ OA-specific collagen subtypes,^14^ and cryptic immuno-epitopes in the matrix or body fluids resulting from collagen degradation.^15–17^ However, the vital relationship between the structural integrity of collagen and OA development remains to be established at the molecular level. Here, we examine and delineate the spatiotemporal changes in the unfolding of collagen molecules in cartilage and surrounding knee joint tissues during early OA progression using a CHP that we developed to specifically bind denatured collagen chains through triple-helical hybridization.^12^ With investigation into its underlying driving forces, we propose that the denatured collagen molecule is a crucial hallmark of early OA damage in the joint, and showcase its advantages in therapeutic evaluation and molecular magnetic resonance imaging for early OA.

## METHODS

This study aims to address the unmet clinical need for defining and noninvasively detecting OA from the pre-osteoarthritic stages. These goals were addressed by (i) interrogating the molecular integrity of the collagen and demonstrating collagen denaturation as the structural hallmark of early OA through histological and in vivo collagen hybridization and ex vivo 3D whole-mount joint imaging in rodents, (ii) investigating the mechanical and biological driving forces of molecular collagen defects in OA cartilage damage, (iii) evaluating the therapeutic efficacy of agents for early OA in vivo in rat models, (iv) developing an MR contrast agent and demonstrating its capacity and specificity in identifying early OA changes in rats, and (v) initiating a pilot study to translate these findings using OA patients’ life-size cartilage specimens. All procedures for animal experiments were reviewed and approved by the Experimental Animal Ethics Committee of the Fifth Affiliated Hospital of Sun Yat-sen University (approval no. 00151). All mice received humane care under the criteria set out in the *Guide for the Care and Use of Laboratory Animals* published by the National Institutes of Health. In all animal experiments, mice of similar age and size were randomly assigned to each group. The investigators were blinded for histological and MRI scoring. Because collagen-hybridizing MRI is a new imaging technology, it is difficult to estimate sample size with adequate power. All in vivo and in vitro experiments were replicated at least three independent times. Images acquired from tissue staining (e.g., figure 1A,B), in vivo or ex vivo fluorescence imaging (e.g., figure 1E, figure 3B, and online supplemental figure S5), and light-sheet fluorescence microscopy (e.g., figure 2, online supplemental figure S7 and S8) are representative of similar results from at least three mice from each group. The details of the study design, sample size, experimental replicates, and statistics are provided in the corresponding figures and figure legends. Human OA tissue samples were from Nanjing Drum Tower Hospital. The Ethics Committee of Nanjing Drum Tower Hospital of Nanjing University approved these studies (approval no. K228-1). Informed consent was obtained from the patients. The online supplementary materials provide details about peptide synthesis and labeling, near-infrared fluorescence imaging, histology and microscopy, MR imaging, and statistical analysis.

**Figure 1.**
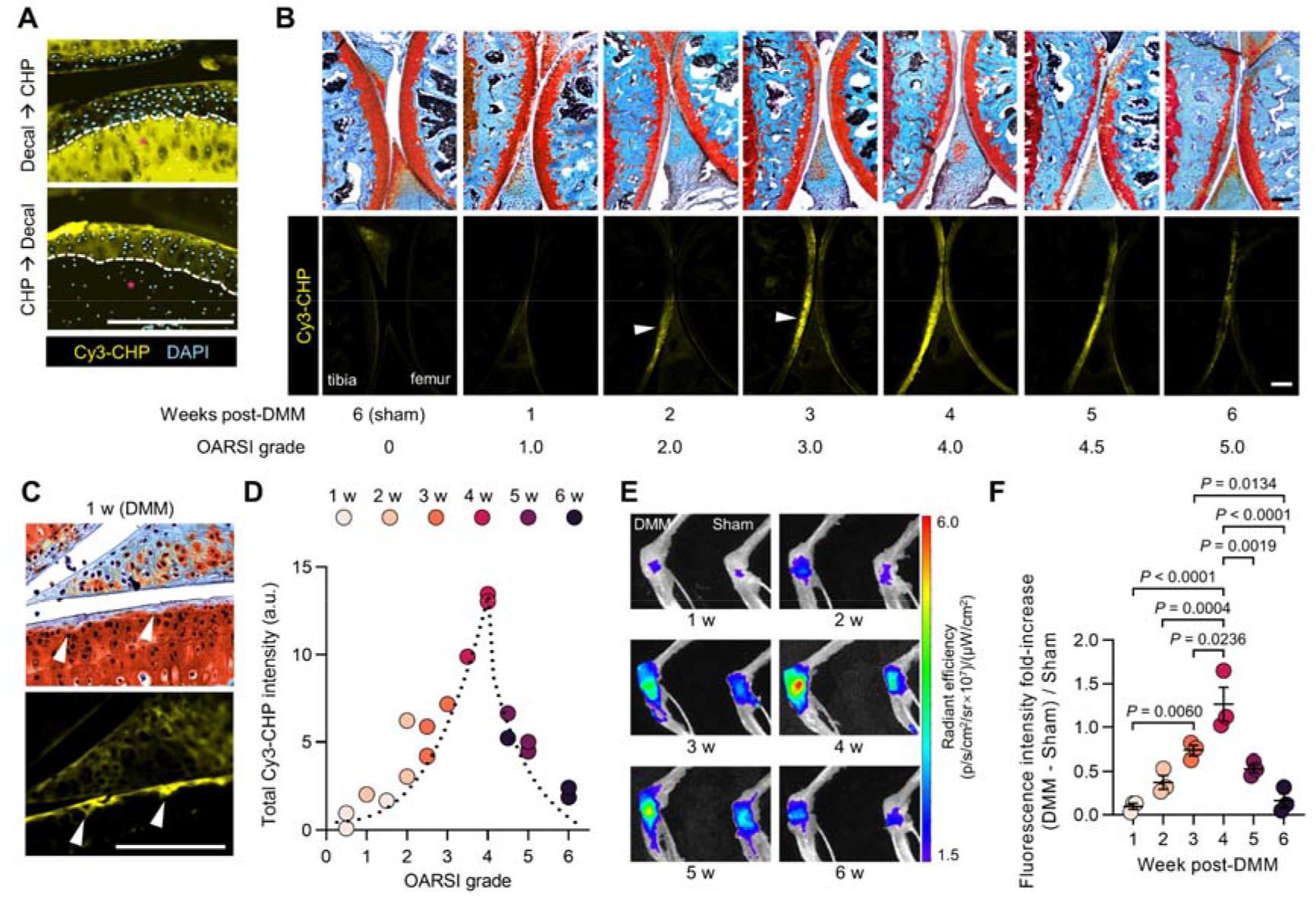
Molecular collagen denaturation in the knee articular cartilage during early osteoarthritis (OA) revealed through histologic and in vivo collagen hybridization. (A) Unlike standard hard tissue processing [decalcification (decal) before CHP staining] that results in extensive artificial collagen denaturation and Cy3-CHP staining (top) in bone (*) in cryosections of normal mouse knee joints, our customized processing (staining before decalcification) leaves the triple-helical folding of collagen molecules intact (bottom, detailed in online supplemental figure S1). (B) Representative images of cryosections from the knee joints of C57BL/6 mice with surgical destabilization of the medial meniscus (DMM, right knee) or sham operation (left knee), harvested weekly post-operation (*n* = 3 mice per week) for histology with Safranin O/fast green (top) and Cy3-CHP (bottom). (C) Up-close histology images of a mouse knee joint one-week post-DMM surgery (OARSI = 0.5) exhibiting overall intact cartilage morphology but with Cy3-CHP fluorescence localizing to micrometer-size surface defects with minor loss in Safranin-O coloration (arrowheads in B, C). (D) Total Cy3-CHP fluorescence intensities quantified from all knee joint sections (images in B and fig. S3), plotted against their corresponding OARSI grades. (E) Representative fluorescence images of the mice’s knee joints collected (3 h post intravenous injection of 1 nmol of Cy5-CHP) weekly post-DMM-operation (*n* = 3 mice per week). (F) Excess Cy5-CHP fluorescence signals from the DMM-injured knee compared to the paired sham-operated knee in individual mice measured each week post-operation. Additional images and data are shown in online supplemental figures S1-S6. Data are mean ± s.e.m. and were statistically analyzed using one-way ANOVA followed by Tukey’s multiple-comparison tests (F). Scale bars: 300 μm (A-C).

**Figure 2.**
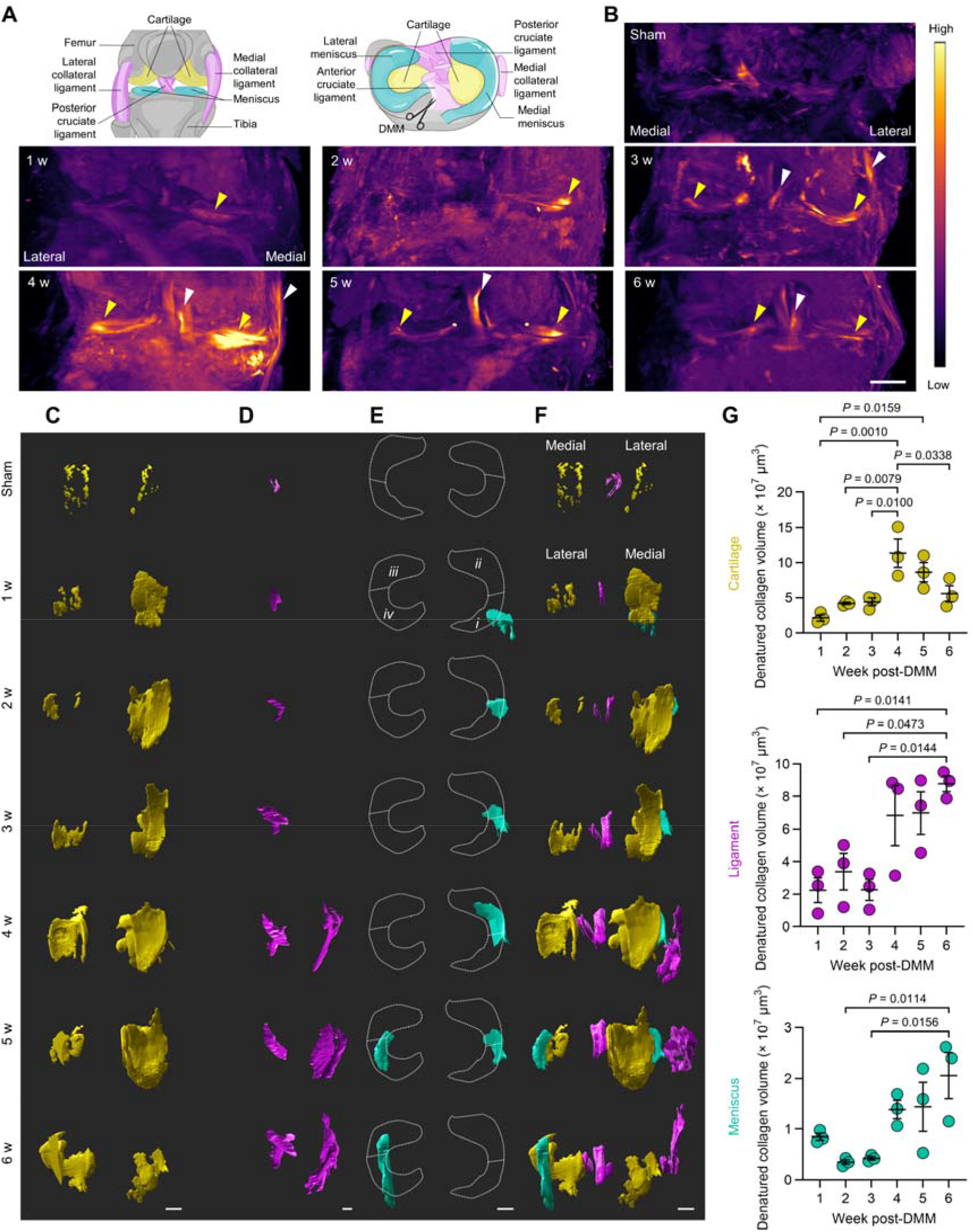
Spatiotemporal landscape of molecular collagen defects in knee joint components during early OA progression. (A) Anatomy of the right knee joint in the anterior and superior views. Scissors indicate the location of DMM surgery. (B) Light-sheet fluorescence microscopy images of the sham-operated mouse knee joints compared to the joints collected 1 to 6 weeks post-DMM-operation. The specimens were harvested for clearing 3 h post-intravenous injection of Cy5-CHP, which was uptaken by the joint tissues in vivo. Arrowheads indicate locations with notable collagen denaturation in cartilage (yellow) and ligaments (white). (C to F) Three-dimensional (3D) reconstruction of the mice’s joints showing Cy5-CHP fluorescence signals from cartilage (C), ligament (D), and meniscus (E, *i*➔*iv*: counterclockwise) overlaid in (F, G), Changes in the lesion volume with molecular collagen defects in the cartilage, ligaments, and meniscus post-DMM operation, measured by the volumes of CHP-positive tissues identified in the 3D fluorescence reconstruction (C to E). Additional images and results are in online supplemental figures S7-S9. Data are mean ± s.e.m. (*n* = 3 mice per time point) and were statistically analyzed using one-way ANOVA followed by Tukey’s multiple-comparison tests (G). Scale bars: 500 μm (B), 300 μm (C-F).

**Figure 3.**
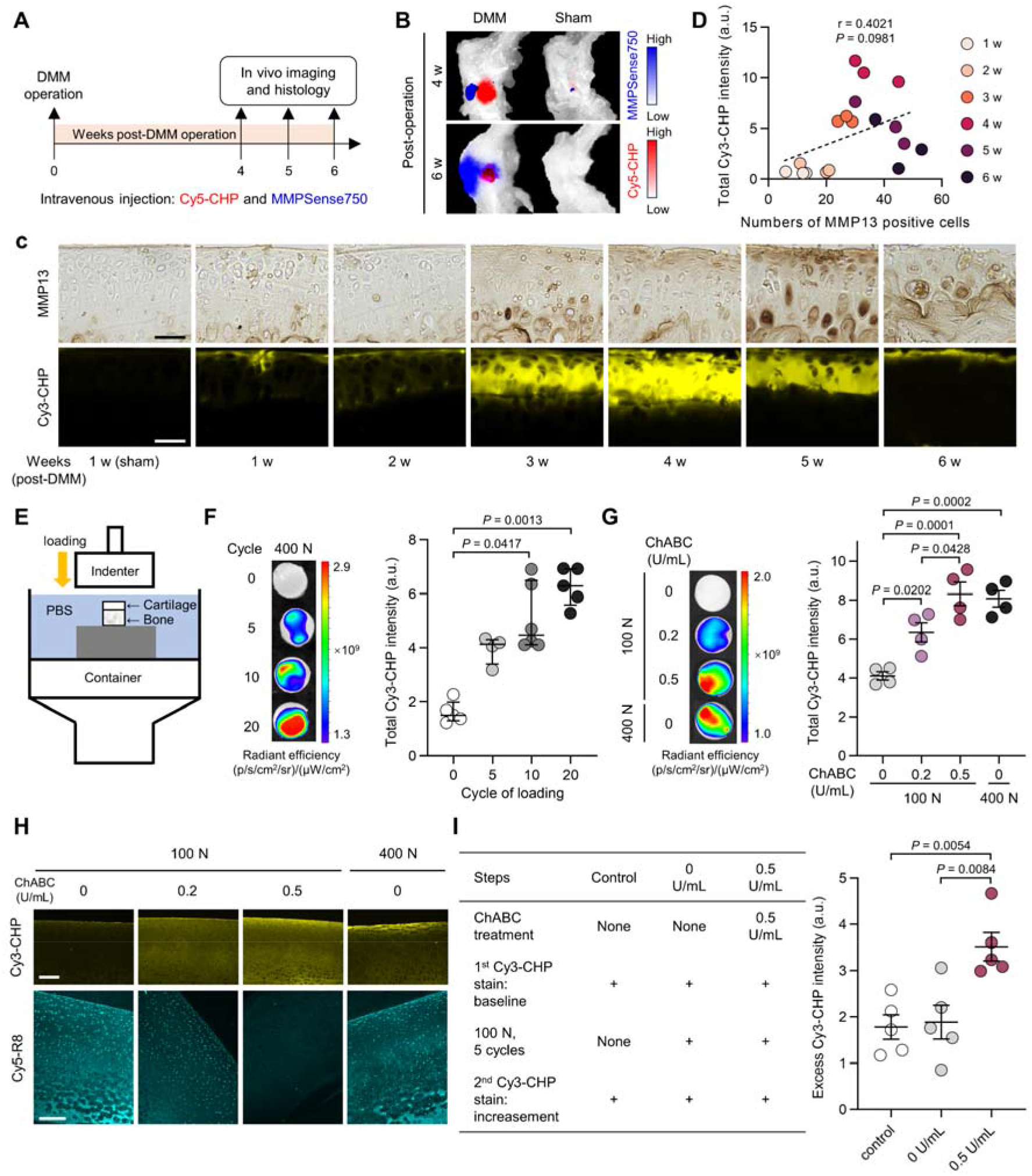
Early collagen denaturation post-DMM-injury is driven by mechanical insults, not MMPs. (A) Schematic of the DMM-injured mice (*n* = 3 mice per time point) administered in vivo with both Cy5-CHP and MMPSense750 for knee joint imaging. (B) Representative up-close ex vivo fluorescence images of the mice’s knee joints 4- and 6-weeks post-operation showing distinct localization of Cy5-CHP (3 h p.i.) and MMPSense750 (24 h p.i.) signals. (C) Paired images of cryosections from the DMM (or sham) operated knee joints harvested weekly post-operation and co-stained with an anti-MMP13 antibody (immunohistochemistry) and Cy3-CHP. (D) Cy3-CHP fluorescence quantified for each cartilage section showed poor correlation with the number of MMP13-positive cells, as interpreted by the Pearson correlation coefficient. (E) Schematic of the setup for the mechanical loading tests on an articular osteochondral plug specimen (diameter: 1 cm) excised from the porcine tibia plateau. (F) Representative fluorescence images (left) and quantification (right) of normal cartilage samples stained with Cy3-CHP after 0-20 cycles of 400 N (i.e., 5.1 MPa) loading. Data: median with interquartile range, statistics: Kruskal-Wallis, Dunn’s. (G) Representative fluorescence images (left) and quantification (right) of the normal or chondroitinase ABC (ChABC) treated cartilage samples after 100 or 400 N loading and staining with Cy3-CHP. (H) Representative fluorescence images of the cryosections of the Cy3-CHP-stained cartilage samples from (G). A neighboring section was stained by a highly cationic peptide Cy5-Ahx-RRRRRRRR (Cy5-R8) to verify the GAG loss due to ChABC treatments. (I) Experimental design and outcome, recorded as the quantified fluorescence increment from sequential Cy3-CHP staining of normal or ChABC-treated porcine cartilage samples before and after 5 cycles of 100 N loading. Additional images and results are in online supplemental figures S10-S12. Data are mean ± s.e.m. Statistical analysis was performed using one-way ANOVA followed by Tukey’s multiple-comparison tests (G and I). Scale bars: 34 μm (C), 250 μm (H).

## RESULTS

### Collagen denaturation in mouse knee cartilage during early OA

We utilized a widely adopted OA model prepared by surgical destabilization of the medial meniscus (DMM) in the right knee of a C57BL/6 mouse paired with a sham operation in the left knee.^18^ ^19^ To examine collagen denaturation in cartilage histologically during OA progression, we harvested knee joints from these mice weekly post-operation and stained them with fluorescently labeled Cy3-CHP in vitro, following a customized decalcification procedure that leaves the folding of collagen molecules intact for CHP staining (figure 1A, online supplemental figure S1 and S2). The cartilage of the DMM joints exhibited only subtle loss of glycosaminoglycan (GAG) without signs of erosion within the first 3 weeks post-operation; yet, there was already notable CHP binding at the cartilage surface, often in locations with mild GAG loss (figure 1B, arrowheads, and online supplemental figure S3) even as early as 1-week post-DMM (figure 1C). The size and fluorescence intensity of CHP-stained cartilage increased steadily during this stage (OARSI grade 0-3, figure 1B,D), with the tibial side showing stronger CHP signals than the femoral side (figure 1B). The CHP binding level and area increased drastically and peaked at OARSI grade 3.5-4 (usually 4 weeks post-DMM, figure 1B,D), when the total CHP fluorescence signal from the cartilage section approximated at 10 times that of the sections of OARSI grade 1 (figure 1D). Beyond 4 weeks post-DMM, substantial cartilage erosion was seen histologically (OARSI grade ≥ 4, figure 1B) with immunostaining showing great loss of collagen II down to deep cartilage layers (online supplemental figure S4). Meanwhile, the area and fluorescence of the CHP-stained cartilage were also diminishing accordingly (figure 1B,D, and online supplemental figure S3).

To verify our findings in vivo, we performed near-infrared fluorescence imaging of the mice weekly post-operation (p.o.) following tail vein injection of a Cy5-labeled CHP. Consistently, the DMM-injured knee exhibited stronger fluorescence from in vivo uptake of Cy5-CHP (but not the sequence-scrambled control peptide Cy5-^S^CHP) than the sham-operated knee of the same mouse each week p.o. (online supplemental figure S5A,B). The DMM-injured knee joints collected 3 h post-CHP-injection showed steadily increasing ex vivo fluorescence until 4 weeks p.o. (figure 1E,F, and online supplemental figure S5C,D) when the CHP uptake peaked in the diseased joints (figure 1F, online supplemental figure S5D). This trend strongly correlated with the above CHP histology findings (figure 1B,D). Furthermore, similar results were obtained in rats post-DMM operation with intra-articular injection of Cy5-CHP (online supplemental figure S6). Overall, these results (figure 1) demonstrate that collagen denaturation may occur much earlier than histomorphologic changes in the DMM-induced OA joints and can be targeted in vivo by CHP hybridization.

### Landscape of molecular collagen defects

Using light-sheet microscopy, we fluorescently imaged the CHP uptake in the DMM mouse knee joints collected 3 h post-intravenous-injection of Cy5-CHP (figure 2A,B, online supplemental figure S7 and S8), and performed 3D reconstruction (figure 2C-F, online supplemental figure S9) and spatial measurements of the molecular lesions with higher-than-normal CHP fluorescence (figure 2G). Such molecular mapping reveals distinct, meaningful spatiotemporal features of collagen defects in major anatomical components during early OA progression. (1) Higher-than-sham CHP fluorescence could be found in the medial cartilage of the DMM joints as early as 1 week p.o. (figure 2B, yellow arrowheads); the fluorescence and volume of the CHP-labeled cartilage lesion (figure 2B,G) peaked at 4 weeks p.o. (figure 2C,G). From 1 to 4∼5 weeks p.o., the molecular cartilage defect was greater on the medial side than the lateral (especially in the tibial plateau, figure 2B,C, online supplemental figure S9A). (2) Collagen denaturation was modest in the anterior and posterior cruciate ligaments 1-3 weeks p.o. (figure 2D, online supplemental figure S9C) but became extensive in the medial collateral ligaments after 4 weeks p.o. (figure 2D) increasing the volume of ligament collagen defects (figure 2G). (3) The region rich in denatured collagen appeared to rotate counterclockwise in the superior view of the post-DMM-surgery meniscus (figure 2E, region *i*➔*vi*, online supplemental figure S9D) with the destabilized medial anterior horn next to the transected ligament first to suffer from collagen denaturation (region *i*).

### Driving forces of collagen denaturation in early post-DMM cartilage

Next, we examined the driving forces of collagen denaturation in OA knee cartilage. Matrix metalloproteinases (MMPs), especially MMP13, are considered the main collagen catabolic factor in OA pathogenesis.^20^ ^21^ We thereby first tested the correlation between MMP activities and collagen denaturation by in vivo imaging the DMM mice administered with both Cy5-CHP and MMPSense750, a fluorescence probe for broad-spectrum MMP activities (figure 3A).^22^ Unexpectedly, the fluorescence of Cy5-CHP and MMPSense750 showed distinct localizations in the same OA knee joints (figure 3B and online supplemental figure S10A). Moreover, the normalized intensity of MMPSense750 from the DMM-injured knees (i.e., fold change in fluorescence of each mouse’s DMM-injured knee over the sham-operated knee) did not increase until 6 weeks post-DMM surgery (online supplemental figure S10B), displaying a temporal profile of in vivo MMP activity distinct from that of Cy5-CHP uptake (online supplemental figure S10B,C). Immunohistochemical staining showed that MMP13 remained non-detectable in the cartilage until 5 weeks p.o. when it became highly concentrated in the superficial zone; however, the Cy3-CHP fluorescence in the same cryosections was localized to the middle and superficial zones during 3-4 weeks p.o., overlapping minimally with the MMP13-positive coloration (figure 3C and online supplemental figure S11A). Also, the number of MMP13-positive cells correlated poorly with the level of Cy3-CHP binding in the same cartilage tissues (figure 3D, online supplemental figure S11B). Overall, the inconsistency between MMP expression and CHP binding across multiple spatiotemporal profiles, suggests that the early cartilage collagen denaturation in the DMM-injured mice may not be adequately explained by MMP degradation.

The above results prompted us to investigate whether and how mechanical insults can cause molecular collagen denaturation in cartilage. We stained osteochondral plug specimens excised from normal porcine patella-femoral joints with Cy3-CHP, which had been compressed in vitro (figure 3E) with increasing levels (online supplemental figure S12A) or cycles (figure 3F). The images and fluorescence quantification indicated that loading caused collagen denaturation that allowed triple-helix specific CHP hybridization (online supplemental figure S12B) and that the denaturation increased with the level of the applied stress and cycle (figure 3F, online supplemental figure S12A,C). Cross-sectional confocal microscopy showed that collagen denaturation was concentrated in the superficial layer of these loaded cartilage tissues to ∼150 μm deep (online supplemental figure S12D,E).

We often noted that whether it is introduced in vivo or ex vivo, CHP binds to cartilage more prominently in locations with even subtle loss of GAGs (e.g., figure 1B,C, online supplemental figure S6D). Considering GAGs’ vital contribution to swelling and compressive resistance in cartilage,^10^ ^23^ we hypothesized that GAG loss intensifies mechanical disruption to collagen molecules. To test this, we treated porcine cartilage samples with chondroitinase ABC (ChABC) to selectively reduce their GAG content before loading.^24^ Post-loading CHP fluorescence stain showed more collagen denaturation with increasing ChABC concentrations under the same loading (figure 3G); for samples treated with 0.5 U/mL ChABC, loading of merely 100 N yielded a degree of collagen denaturation that would require loading of 400 N on normal untreated samples (figure 3G and online supplemental figure S12F). Cross-sectional fluorescence images verified that the same loading caused increasing collagen denaturation within deeper zones in cartilage with declining GAG content (figure 3H).^25^ Finally, successive CHP staining before and after loading revealed that a mild process (100 N, 5 cycles) that induced no collagen unfolding to normal, GAG-rich cartilage resulted in substantial collagen denaturation and CHP fluorescence increment in samples with GAG removal (figure 3I). These results demonstrated GAGs’ protection to collagen molecules against mechanical denaturation in cartilage. All combined (figure 3), the data suggest that the molecular integrity of collagen in cartilage can be compromised by mechanical factors during early OA progression.

### Therapeutic assessment for early OA

To showcase the potential of using collagen denaturation in therapeutic assessment for early OA, we live imaged the knee joints of DMM-operated rats weekly with intra-articularly dosed Cy5-CHP to monitor two treatments in vivo: oral glucosamine sulfate (GlcN) and intra-articular hyaluronic acid (HA) (figure 4A). Both treatments have shown preclinical efficacy against OA development.^26–29^ According to the in vivo Cy5-CHP fluorescence, both treatments reduced collagen denaturation within the DMM joints at 4-7 weeks p.o., keeping the CHP signal at a minimal level approximating the sham-operated knees in individual rats (figure 4B,C, online supplemental figure S13). At 7 weeks p.o., the in vivo CHP uptake in the DMM-knee was significantly lower in the two treatment groups than in the untreated group (figure 4C). The therapies’ attenuating effect on cartilage destruction was further corroborated by the endpoint histopathology (figure 4D and online supplemental figure S14) and in situ fluorescence examination of the in vivo Cy5-CHP uptake in the cartilage tissue (figure 4E).

**Figure 4.**
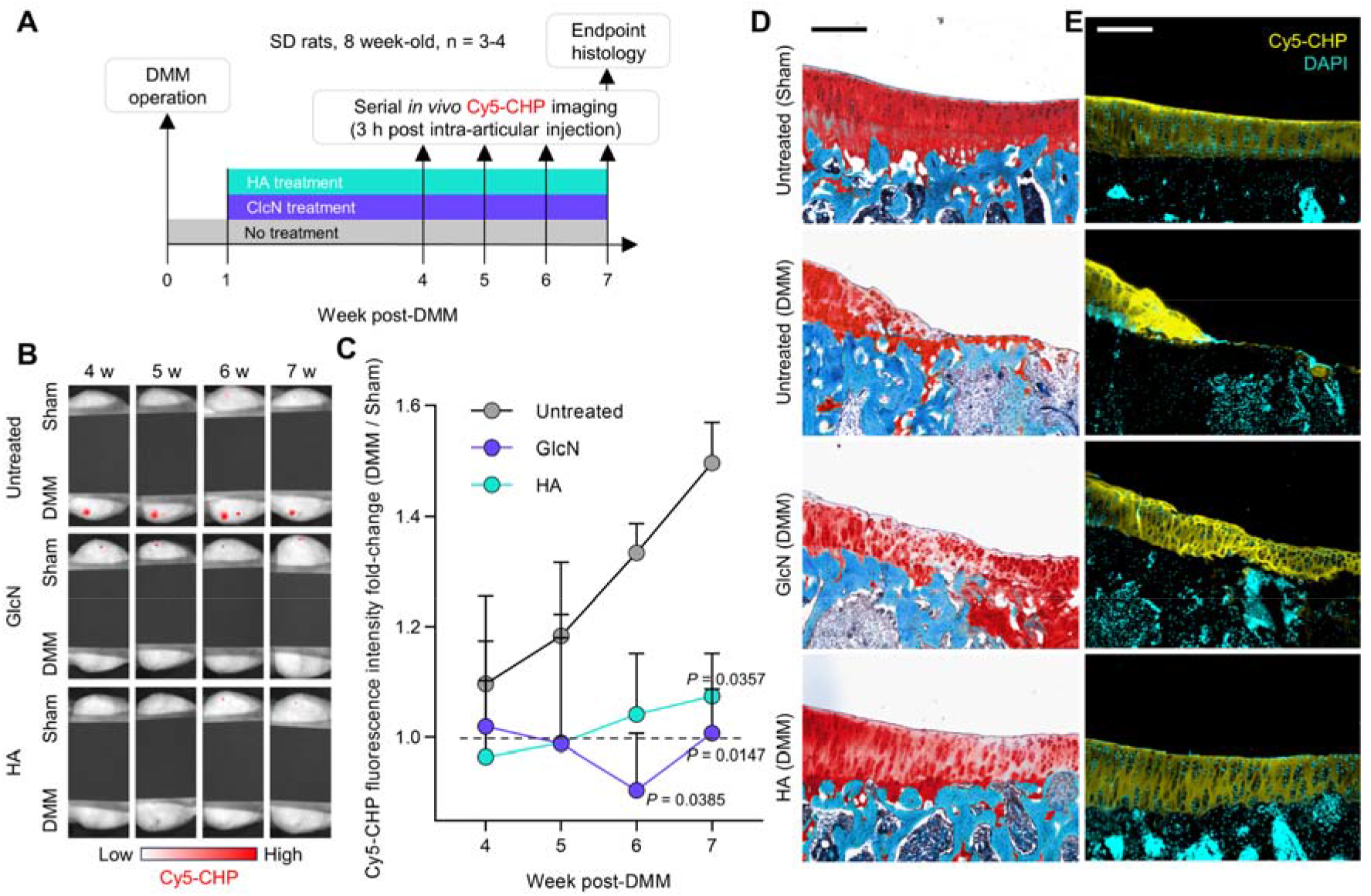
Evaluation of treatments for early OA by collagen hybridization. (A) DMM-operated SD rats (*n* = 3–4 mice per group) were treated with glucosamine sulfate (GlcN, daily, oral) or hyaluronic acid (HA, weekly, intra-articular injection) and monitored longitudinally through in vivo Cy5-CHP fluorescence imaging. (B) Representative in vivo fluorescence images obtained 4–7 weeks post-operation showing minimal Cy5-CHP uptake in each DMM-operated knee joint for the GlcN and HA groups compared to the untreated DMM group. The fluorescence display of each image has been individually adjusted such that a minimal intensity is shown for the sham-operated knee. (C) Serial measurements of the ratio of the in vivo Cy5-CHP fluorescence signals between the DMM- and sham-operated knee joints of the rats in each group. (D and E) Representative micrographs of endpoint histology at 7 weeks post-operation showing the knee cartilage integrity of the rats from each group (D, Safranin O stain) and the matching fluorescence (E) due to the in vivo cartilage uptake of Cy5-CHP (harvested 3 h post-CHP-injection). The tibial plateau cartilage of the sham knee was intact with low CHP binding to the surface; for the GlcN-or HA-treated DMM cartilage, minor GAG loss with modest surface irregularity and fibrillation was found in locations with moderate CHP fluorescence. In contrast, the cartilage of the untreated DMM joints showed severe defects extending to the subchondral bone with substantial CHP uptake in the remaining surrounding cartilage. Additional images are in online supplemental figure S13 and S14. Data are mean ± s.e.m. and analyzed using repeated measures two-way ANOVA followed by Tukey’s multiple-comparison tests (C). Scale bars: 250 μm (D and E).

### In vivo magnetic resonance imaging

To enable in vivo detection of molecular collagen defects in the joints by MRI, we synthesized Gd_n_-Cy5-CHP by conjugating Cy5-CHP with multiple Gd-ion chelates, creating a contrast agent with sensitivity severalfold that of Magnevist, a common clinical MR contrast agent (figure 5A, detailed in online supplemental figure S15 and methods). Compared to the pre-contrast scans, intra-articularly administered Gd_n_-Cy5-CHP markedly enhanced the T1 signals in the tibia cartilage and ligaments of the DMM-injured (as opposed to the sham-operated) rat knee joints (figure 5B,C). The high signal sites in the cartilage shown in T1WI imaging matched closely with the postmortal fluorescence results of the isolated joints, both indicating Gd_n_-Cy5-CHP’s in vivo binding to cartilage locations with GAG loss (figure 5D,E). Furthermore, longitudinal Gd_n_-Cy5-CHP enhanced T1WI scans of the same individuals depicted changes in the level and location of collagen defects in cartilage and ligaments over weeks during early OA progression (figure 5F); these subtle damages were not readily visible in the T2WI scan routinely used for clinical examination of soft tissue degeneration (figure 5D,F).

**Figure 5.**
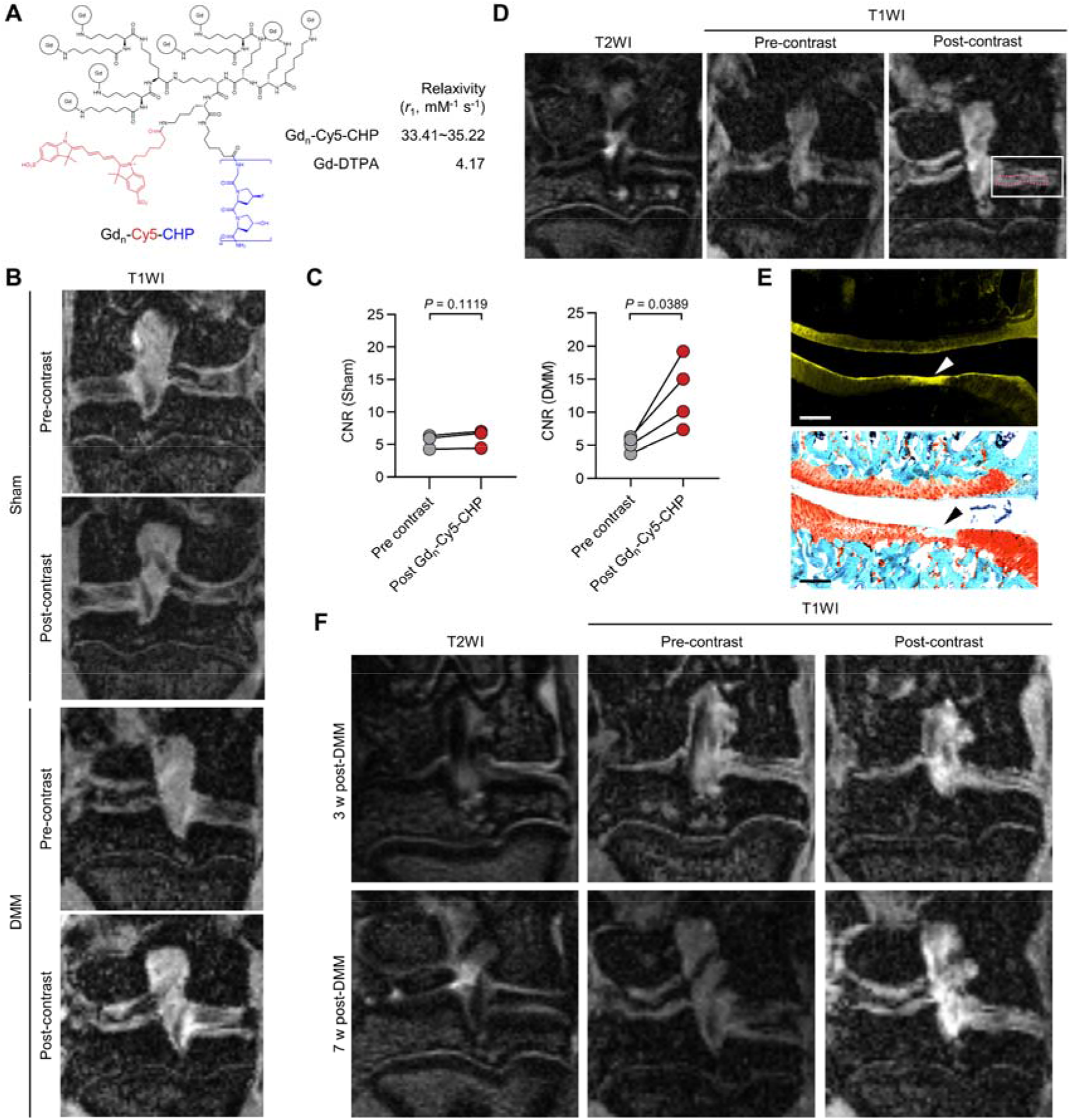
In vivo magnetic resonance (MR) imaging of molecular collagen defects in DMM-injured rat knee joints. (A) Chemical structure of contrast agent Gd_n_-Cy5-CHP and its measured longitudinal relaxivity compared to Gd-DTPA (Magnevist). (B) Representative in vivo T1WI images of the sham- or DMM-operated knee joints of the same rat 7 weeks post-surgery. The rats were scanned before and after intra-articular injection of 250 nmol of Gd_n_-Cy5-CHP. (C) Contrast-to-noise ratio (CNR, detailed in online supplementary materials) quantified from regions of interest in the medial tibial plateau cartilage of the knee pairs before and after contrast enhancement in individual rats imaged in c (*n* = 4, statistically analyzed using paired t-test). (D) Representative in vivo T1WI images of the DMM-injured rat knee joints 7 weeks post-operation pre- and post-contrast enhancement with Gd_n_-Cy5-CHP, compared to the T2WI image of the same joint. (E) Images of the Safranin O stain (bottom) and in situ fluorescence (top) of the sections from the cartilage within the boxed region in (D), highlighting Gd_n_-Cy5-CHP’s selective binding to spots with GAG loss in vivo. (F) Repeated in vivo T1WI (with or without Gd_n_-Cy5-CHP contrast enhancement) and T2WI scans of the same DMM-injured rat knee joint 3- and 7-week post-operation. Additional data in online supplemental figure S15. Scale bars: 250 μm (E).

### Ex vivo study of clinical cartilage tissues

We prepared osteochondral plug specimens from different regions of femoral cartilage surgically removed from OA patients during knee replacement (figure 6A). We grouped these samples into three [group i: mild disruption (OARSI grade ≤ 2), ii: moderate degeneration (2 < OARSI grade ≤ 4), and iii: severe damage (OARSI grade > 4)] based on the cartilage morphology (Fig. 6A) and histopathology (figure 6B) and labeled them with Gd_n_-Cy5-CHP ex vivo. The T1WI magnetic resonance and fluorescence imaging showed that the cartilage plugs in group ii exhibited the most pronounced CHP signals near the less-smooth cartilage surface (figure 6C-E, online supplemental figure S16A,B). In situ fluorescence and histology from the post-MRI cryosections revealed that Gd_n_-Cy5-CHP accumulated strongly to regions of significant GAG loss close to the cartilage surface in group ii while being less effective in targeting the cartilage in group i in which collagen was not yet disrupted and in group iii, where collagen had been substantially eroded (figure 6F and online supplemental figure S16). Three-dimensional light-sheet fluorescence imaging of CHP-labeled, bulk knee cartilage from OA patients delineated the heterogeneous spatial distribution of denatured collagen in the tissue (figure 6G, online supplemental figure S17). The above results are highly consistent with our findings in animal models (figures 1-5), suggesting that CHP can effectively detect molecular collagen defects in the modestly damaged cartilage of OA patients.

**Figure 6.**
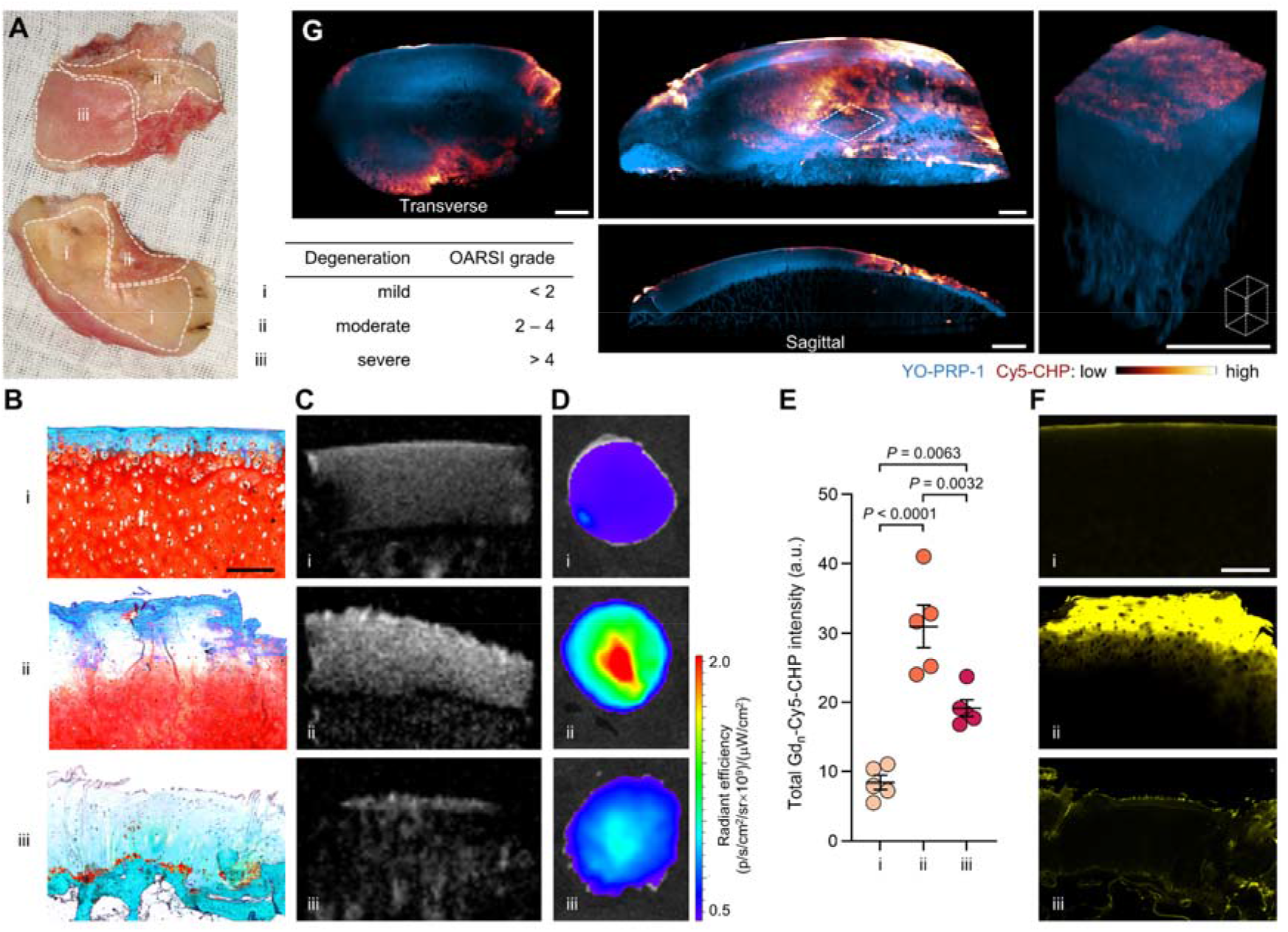
Ex vivo identification of molecular collagen defects in degenerated cartilage from OA patients. (A) Photograph of representative femoral cartilage specimens surgically removed from OA patients during knee replacement. i-iii: Regions with mild-to-severe levels of cartilage degeneration and OARSI grades. Osteochondral plug specimens were excised from regions i-iii (*n* = 5 plugs per group, diameter: 6 mm). (B to D) Representative images of the histology (B, Safranin O stain), T1WI magnetic resonance (C), and fluorescence imaging (D) of the plug specimens labeled with Gd_n_-Cy5-CHP ex vivo. (E) Quantification of the fluorescence intensity from Gd_n_-Cy5-CHP in the cartilage samples of groups i-iii. (F) In situ Gd_n_-Cy5-CHP fluorescence in cryosections adjacent to the ones in (B), cut from the plug specimens of groups i-iii. (F) Three-dimensional light-sheet fluorescence scans of a life-size femoral knee cartilage specimen from an OA patient labeled with Cy5-CHP. Cell nuclei were stained with YO-PRO-1. The boxed area is zoomed in on the right. Data are mean ± s.e.m. and statistical analysis was performed using one-way ANOVA followed by Tukey’s multiple-comparison test (E). Additional images in online supplemental figure S16 and S17. Scale bars: 250 μm (B and F), 3 mm (G).

## DISCUSSION

OA typically develops over decades, presenting a window of opportunity for intervention. A major undertaking in current OA management is to transition from late disease alleviation toward prevention.^3^ ^7^ ^8^ To empower and accelerate this paradigm shift, the pre-osteoarthritic stages of joint degeneration must be defined with new biomarker discovery and diagnosis development.^8^ ^30^ ^31^ Therefore, we interrogated the integrity of the most basic building block of joint anatomy - the molecular folding of the collagen architecture - from OA onset and established collagen denaturation as a disease hallmark preceding collagen proteolysis (figure 3, online supplemental figure S10 and S11) and loss (figure S4) that result in apparent histopathologic and radiographic changes (OARSI grade 1-4; figure 1 and 6).^32^ We further mapped and measured the molecular lesions three-dimensionally in full joint anatomy along the course of OA progression in mice (figure 2), delineating the molecular landscape of collagen destruction in early OA pathogenesis and biomechanics.^33^

This study exemplifies a new approach to characterizing tissue degeneration from the perspective of material science^34^ and structural biology. The collagen matrix is otherwise equally abundant in normal and pre-osteoarthritic joints and therefore can be overlooked in search of molecular biomarkers, yet it may be the first to suffer from structural alteration in degeneration and injury. Herein, the disease characteristic arises from the abnormality in protein structure instead of quantity, offering a new category of biomarkers not easily identified by the conventional ‘candidate protein’ approach.

Investigating the mechanisms driving early-stage OA progression is crucial for developing effective treatments and risk management.^35^ We identified that mechanical overload, more than protease-degradation, is an underlying driving force of molecular collagen denaturation in early cartilage degeneration (figure 3); we further showed that the compressive-resistant GAG component plays a decisive protective role against the mechanical collagen denaturation in cartilage (figure 3).^36–38^ These findings provide molecular structural evidence that closes the gap in early OA biomechanics and pathophysiology and underlines weight, joint overloading, and GAG loss as modifiable risk factors to reduce the disease burden.^35^ ^39^ ^40^ Furthermore, GAG loss is one of the first signs of early OA but is challenging to image and target.^13^ ^41^ Our study uncovered a strong spatiotemporal and logical association between collagen denaturation and GAG loss (figure 1B,C, figure 3G-I, figure 6, online supplemental figure S6D and S16), providing an indirect strategy for detecting and targeting GAG loss within the degenerating cartilage.

To date, no validated diagnostic criteria are available for early-stage knee OA.^8^ The detection, monitoring, and therapy development of early OA have been hampered by the insensitivity of the conventional radiographic measures of joint health.^7^ We demonstrated that the CHP (even intravenously administered) can target the denatured collagen and deliver diagnostic modalities to the avascular articular cartilage in vivo (figures 1-3),^42^ ^43^ allowing straightforward monitoring of knee joint integrity and therapeutic assessments through live imaging (figure 4). Such noninvasive detection of early joint defects will facilitate the development of new disease-modifying therapies targeting the pre-osteoarthritic condition^44^ and reduce the reliance on post-mortal histology. MRI is currently the most common modality for direct imaging of soft tissues in the clinic and research.^45^ ^46^ As such, we developed contrast agent Gd_n_-Cy5-CHP and showcased its sensitivity by highlighting the subtle cartilage defects beyond the capacity of routine T2-based morphological assessment (figures 5). We further demonstrated that the MR-fluorescence CHP agent can visualize molecular collagen defects in human knee cartilage with only mild-to-moderate degeneration (i.e., OARIS grades 1-4) and depict the defect heterogeneity in 3D over life-size patient specimens (figures 6). OA is a heterogeneous disease; such precision in illustrating collagen defects may contribute to the much-desired molecular stratification of early OA population.^47–49^ Overall, these translational findings set the stage for further clinical development of collagen hybridization in the early diagnosis and treatment of OA among a range of musculoskeletal degenerative diseases and injuries.

## Acknowledgments

We thank Profs. Jiang Xia, Chao Liu, and Hang Lin for the informative discussion, Dr. Xianghe Xu for the guidance in producing the DMM model, and Yuqi Lin for assistance during MR imaging. Online supplemental figure S2 was created with BioRender.com.

## Contributors

Conceptualization: YL, DS. Methodology: HK, YL, RQ, YF, XL, ZL, TJ. Investigation: HK, RQ, YF, DZ, XL, ZL, TJ, YXF (Yuxiang Fei), YH. Visualization: HK, RQ, YF, DZ, DNZ (Daoning Zhou), WW, YZ, YL. Funding acquisition: YL, DS. Project administration: DS, YL. Supervision: SL, DS, YL. Writing—original draft: HK, RQ, DZ, YF, XL, YL.Writing—review & editing: YL, DS.

## Ethics approval

In the study, all animal experiments were approved by the experimental animal use and ethics committee of the Fifth Affiliated Hospital of Sun Yat-sen University (protocol number: 00151), and all experimental plans involving knee specimens from patients were approved by the Ethics Committee of Nanjing Drum Tower Hospital (number: K228-1).

## Data availability statement

All data are available in the main text or the supplementary materials. The online supplementary materials include supplementary materials and methods, supplementary table S1, supplementary figures S1-S17, and supplementary references.

