## Supplementary materials for "Targeting Molecular Collagen Defects from the Initiation of Knee Osteoarthritis"


**This PDF file includes:**


Supplementary materials and methods
Supplementary table S1
Supplementary figures S1-S17
Supplementary references (1-13)

### Supplementary Materials and Methods

#### Peptide synthesis and labeling

Cy3-CHP with a sequence of $(GPO)_9$ [O: (2S,4R)-4-hydroxyproline] was used in all histologic tissue staining. Because the $(GPO)_9$ peptide tends to self-trimerize into a triple-helix, it must be heated to 85°C to dissociate the peptide homotrimer into single strands before staining. To avoid this pre-heating step that complicates the tail vein injections, we used Cy5-CHP with the sequence of $(GfO)_9$ [f: (2S,4S)-4-fluoro-proline] within the in vivo, ex vivo, and light-sheet fluorescence imaging studies. This $(GfO)_9$-peptide cannot self-trimerize at body temperature but binds to denatured collagen.[1]

All peptides were synthesized on a Rink Amide-AM resin (loading: 0.37 mmol/g) using the Fmoc/tBu strategy on a PurePep Chorus peptide synthesizer.[2] Fmoc-amino acids were purchased from GL Biochemical. Fmoc-deprotection was carried out by 20% v/v piperidine in DMF for 1.5 min at 50 °C twice. Each amino acid was coupled using HATU (Aladdin), HOAT (Macklin), and DIEA (Macklin) in DMF (resin: amino acid: HATU: HOAT: DIEA = 1: 5: 5: 5: 10) for 5 min at 50 °C twice. Peptides were fluorescently labeled on resin by treating the resin with DIEA (5.0 equiv.) and sulfo-Cyanine3 NHS ester (Cy3, 0.5 equiv.) or sulfo-Cyanine5 NHS ester (Cy5, 0.5 equiv.) for over 24 h in DMSO. Cleavage was performed with a mixture of trifluoroacetic acid (TFA, Macklin) / triisopropylhydrosilane (TIS, Macklin) / water (95:2.5:2.5) for 3 hours at room temperature. The crude peptides were precipitated from the TFA solution by excess cold diethyl ether. The solid was centrifuged, and the supernatant was decanted.

All peptides were purified by reverse-phase high-performance liquid chromatography (RP-HPLC) on a semi-preparative column (Agilent ZORBAX StableBond 300 C18) using a linear gradient mixture of water (0.1% TFA) and acetonitrile (0.1% TFA) (5−35% acetonitrile in 30 min) as the mobile phase. The column was heated to 45 °C in a column oven for $(GfO)_9$ or 80 °C for $(GPO)_9$. The purified peptides were lyophilized and verified by MALDI-TOF on a Shimadzu MALDI-8020 instrument. The freeze-dried peptides were reconstituted in Milli-Q water and the concentrations of peptide solutions were determined using absorbance at 548 nm for Cy3-tagged peptides (extinction coefficient: 162,000 $M^{-1}$ $cm^{-1}$) or 646 nm for the Cy5-tagged peptides (extinction coefficient: 271,000 $M^{-1}cm^{-1}$).

#### Synthesis of MR probes

The magnetic resonance probes $Gd_n$-Cy5-CHP and $Gd_n$-Cy5-$^S$CHP were prepared by first synthesizing the Cy5-labeled sequence $[NH_2\text{-}(AhxK)_2K]_2$KK(Cy5)-Ahx-$(GfO)_9$ on resin before TFA cleavage and HPLC purification (linear gradient: 5−35% acetonitrile in 30 min). The lyophilized Cy5- labeled peptide (94.6 mg, 19.7 μmol), p-SCN-Bn-DOTA (542.4 mg, 788.3 μmol), and DIEA (651 μL, 3942 μmol) were dissolved in anhydrous DMSO (2 mL), and the reaction mixture was stirred at room temperature for 24 h in dark. Subsequently, the DOTA-conjugated peptide was purified by HPLC (linear gradient: 5−50% acetonitrile in 30 min) and lyophilized. The DOTA-peptide conjugate (184.5 mg, 21.8 μmol) and $GdCl_3$ (68.8 mg, 261.0 μmol) were dissolved in 2.5 M sodium acetate buffer (25 mL, pH = 6.4) and stirred at 60 °C overnight. Excess $Gd^{3+}$ ions were removed by Vivaspin 20 (MWCO 3000) through repeated centrifugation at 3000 rpm until no $Gd^{3+}$ ions were detected in the filtrate with xylenol orange.[3-5] The synthesis is schematically described in online supplemental figure S15A. The concentrations of $Gd^{3+}$ ion in the MR probe solutions were determined by inductively coupled plasma optical emission spectroscopy (ICP-OES, Agilent 5800).

**Rodents**

All animal experiments were approved by the experimental animal use and ethics committee of the Fifth Affiliated Hospital of Sun Yat-sen University (protocol number: 00151) and carried out following the regulation requirements. All mice and rats were purchased from the Guangdong Medical Laboratory Animal Center. All animals were housed in ventilated cages with corn cob bedding and provided with reverse osmosis water, a standard diet, and a 12-hour light/dark cycle. Surgical destabilization of the medial meniscus (DMM) was performed on 8-week-old male C57BL/6J mice or Sprague-Dawley (SD) rats to induce OA as reported.[6 7] Briefly, with general anesthesia (2% isoflurane in oxygen), the right knee joint was exposed using a medial parapatellar incision, and the joint capsule was opened gently. The anteromedial meniscotibial ligament in mice was transected using micro-scissors. The left leg of each rat or mouse underwent a sham operation in which only the skin and joint capsule were opened without dissecting the ligaments and removing the meniscus. Finally, the knee joint capsule and skin were closed with a 6-0 silk suture.

**Near-infrared fluorescence imaging**

Each mouse was injected with Cy5-CHP or Cy5-$^{S}$CHP (1 nmol in 100 μL PBS) via the tail vein, 3 h before near-infrared fluorescence imaging with an IVIS Spectrum imager (PerkinElmer Lumina III). MMPSense 750 (NEV10126, PerkinElmer, 2 nmol per mouse) was injected via a tail vein 24 h before NIRF imaging. Hair on the knees was removed before fluorescence imaging. For each SD rat, Cy5-CHP (1 nmol in 50 μL of PBS) was intra-articularly injected 3 h before fluorescence imaging. The animals were usually sacrificed and their leg specimens were collected, imaged, and used in subsequent histology analysis. All fluorescence images were analyzed using the Living Image software (PerkinElmer). The total fluorescence signal from each knee was measured by quantifying the radiation efficiency within the selected circular region of interest of consistent sizes in the images. In addition, CHP-stained porcine and human knee cartilage specimens were also performed fluorescence imaging and quantitative fluorescence signal analysis as described above.

**Histology and microscopy**

***Tissue processing***

All paraffin-embedded sections of the knee joints were prepared following standard procedures by Servicebio Technology. To prepare the cryosections for Safranin O staining or fluorescence microscopic examination of in-vivo-bound Cy5-CHP, rodent knee specimens were fixed in 4% paraformaldehyde (PFA) at room temperature overnight and washed with PBS three times. The fixed specimens were decalcified by incubation in a 20% EDTA solution (w/v, pH = 7.0) for one week (mice) or two weeks (rats) with the EDTA solution renewed halfway. After rinsing for 15 min three times, decalcified samples were treated with 30% w/v sucrose solution for 24 h to dehydrate; the samples were immersed in a 30%/40%/30% mixture of sucrose/water/optimal cutting temperature compound (OCT, Solarbio, 4583) for 12 h before frozen in OCT for cryosection. To image the in-vivo-bound Cy5-CHP (e.g., online supplemental figure S6D), the decalcified, dehydrated rat knee tissue was cryosectioned to 10 μm thick. The cryosectioned slides were rinsed with PBS and stained with DAPI (Beyotime C1002, 1:1000 dilution in PBS) for 20 min, then sealed in an antifade mounting medium (Vector, H-1000) and imaged with an EVOS M7000 imaging system.

***Cy3-CHP staining of the knee joint specimens before decalcification***

Each week post-DMM operation, the mouse knee joints were collected and fixed with 4% PFA at room temperature overnight. The fixed knee specimens were cut in half along the median sagittal plane to expose the entire cartilage to the Cy3-CHP solution. Before staining, a Cy3-CHP solution (5 μM in 2 mL PBS) was heated to 80 °C for 5 min to dissociate the triple-helix into single strands and rapidly cooled to room temperature by sitting in an ice-water bath for 2 min. The specimens were incubated in Cy3-CHP solution at 4 °C for 48 h before being washed with 15 mL PBS for 30 min three times. Subsequently, the stained specimens were decalcified with 20% EDTA for 1 week, washed, dehydrated, and cryosectioned to 10 μm thick as mentioned above. The procedure is schematically described in online supplemental figure S1D. The slides were incubated in PBS for 5 min to remove the OCT and sealed in an antifade mounting medium (Vector, H-1000) before microscopic imaging.

***Histology, immunofluorescence, and immunohistochemistry***

Paraffin-embedded tissues were sliced to 5 μm thick using a microtome. Paraffin was removed by xylene, 100% ethanol, 95% ethanol, 85% ethanol, 75% ethanol, 50% ethanol, and PBS for two 5-minute cycles of each solvent. For histological staining, slides of the mouse and rat knee joints were stained with Safranin-O/fast green (Solarbio, G1371) following the manufacturer's instructions for both cryosections or paraffin-embedded sections. The severity of osteoarthritic phenotype was analyzed by two blind independent researchers using the Osteoarthritis Research Society International (OARSI) grades based on cartilage area, synovitis score, osteophyte size, and osteophyte maturity. To detect MMP13, cryosections were used and stained with an anti-MMP13 antibody (Proteintech, 18165-1-AP, 1:300) overnight at 4 °C. To detect collagen II, paraffin-embed sections were deparaffinized and underwent a heat-mediated antigen retrieval process in a pressure cooker at 100 °C for 10-15 min in a sodium citrate buffer (MXB biotechnologies, MVS-0066, pH: 6.0). Subsequently, the sections were stained with a primary antibody against collagen II (Abcam, ab34712, 1:400, online supplemental figure S4A) overnight at 4 °C. For immunohistochemistry, all primary antibodies were detected with a horseradish peroxidase-conjugated secondary antibody at room temperature for color development with DAB (MXB Biotechnologies, DAB-0031) as a chromogen. For immunofluorescence detection of collagen II (online supplemental figure S4B), deparaffinized antigen-retrieved sections were stained with an anti-COL2A1 antibody (ABclonal, A19308, 1:100) followed by an AlexaFluoro647-labeled secondary antibody (goat anti-rabbit IgG H&L, Abcam, ab150079, 5 μg/mL in PBS). The stained slides were imaged with an EVOS M7000 microscope.

***Fluorescence microscopy***

All tissue sections were imaged using an EVOS M7000 imaging system (Thermo Fisher) with 4×, 10×, 20×, or 40× objective lenses and DAPI, RFP, and Cy5 light cubes. The slides stained with Safranin-O/fast green and immunohistochemistry were imaged in a bright field. Large full-section images were generated through view-to-view scanning and image stitching performed automatically by the EVOS imaging system. Pseudo colors were assigned to the images using the LUT color scheme available in the ImageJ software.

**Tissue clearing and light-sheet fluorescence microscopy (LSFM)**

After injection of Cy5-CHP or $Gd_n$-Cy5-CHP and in vivo fluorescence imaging or magnetic resonance imaging, the mice or rats were transcardiacally perfused with 0.02% heparin in PBS (m/v) and 4% PFA to remove blood. Next, the knee samples were collected and cleared following the PEGASOS method.[8] After being fixed in 4% PFA at room temperature for 24 h, the samples were decalcified in 20% EDTA (pH 7.0) solution at room temperature for 1 week (for mouse) or 2 weeks (for rat). The samples were then washed with PBS three times (20 min each round) before being decolorized with 25% Quadrol (Sigma-Aldrich 122262) for 2 days (for mice) or 5 days (for rats). After being washed three times with PBS (1 h each round), the samples were delipidated with tert-butanol (Sigma-Aldrich 360538) and dehydrated with 70% v/v tert-butanol, 27% v/v PEGMEMA500 (Sigma-Aldrich 447943), and 3% w/v Quadrol for 2 days. Finally, the samples were immersed in a clearing medium (BB-PEG) made by 75% v/v benzyl benzoate (Sigma-Aldrich W213802), 22% v/v PEGMEMA500, and 3% w/v Quadrol for at least 1 day. Care was taken to avoid light during the entire clearing process.

Each cleared knee specimen was imaged on a LaVision Biotec Ultramicroscope II equipped with an sCMOS camera. The tissue was immersed in the imaging chamber filled with the BB-PEG medium. Each cleared specimen was scanned with a magnification of 4 on both sides, each side composed of three light-sheet beams with a 5 μm step in the Z-axis. The images were acquired by continuous light-sheet scanning, and stitched using a blend algorithm on both sides. Images were acquired by ImSpector (LaVision BioTec), saved as 16-bit grayscale TIFF images for each channel, and reconstructed with the Imaris software.

**Mechanical loading**

Porcine osteochondral plugs were freshly harvested from the articular cartilage of an adult pig's knee joints obtained from a local slaughterhouse. Plugs (diameter: 1 cm) were excised using an osteotomy drill with constant PBS rinsing for cooling.[9-11] The LTM electrodynamic testing machine was used to apply mechanical loading. The setup consists of a sample holder (filled with PBS) and a vertical rail that provides rigid support for the linear moveable parts. A vertical rod, placed in a stainless-steel housing, performs the mechanical perturbation. After being equilibrated in the holder for 30 min, the samples were loaded in ten loading cycles of 0, 100, 200, or 400 N at a rate of 10 mm/min. For the compression of 200 and 400 N, samples were loaded with 0, 5, 10, or 20 loading cycles respectively. After mechanical loading, the plugs were fixed in 4% PFA at room temperature for 48 h and washed with PBS for 10 min three times.

These fixed plugs were incubated in 5 μM Cy3-CHP or Cy3-$^{S}$CHP solution at 4 °C for 48 h. After being washed with PBS for 2 h three times, the cartilage was removed from the subchondral bone. The plugs were imaged fluorescently and quantitatively analyzed. Moreover, the Cy3-CHP-stained cartilage was cut into 3 mm thick pieces from the articular surface and scanned with a confocal laser scanning microscopy on a coverslip.

All solutions used to treat the unfixed samples in the following steps contained Penicillin and streptomycin (Solarbio, P1400) to prevent bacterial growth. A group of porcine osteochondral plugs was incubated in chondroitinase ABC (ChABC, Sigma, C3667, 0.5 mL per sample) solutions of specific concentrations for 12 h at 37 °C before being washed three times with PBS. Mechanical loading was applied at a 10 mm/min rate while the samples were loaded to 100 N or 400 N for 10 cycles. After mechanical loading, the samples were fixed in 4% PFA for 24 h and washed with PBS.

The samples were stained with Cy3-CHP (5 μM, 1 mL per sample) at 4 °C for 72 h with shaking before extensive washing in PBS. After staining, the plugs were imaged fluorescently with an IVIS instrument with quantitative analysis. The samples were cut into 5 μm-thick cryosections, of which the GAG content was detected using a Cy5-$R_8$ peptide (10 nM, 100 μL per sample, 10 min staining); the Cy3-CHP fluorescence signals in these cartilage sections were also recorded with an EVOS M7000 microscope (figure 3H). For the experiment described in figure 3I, after treating a group of porcine osteochondral plugs in ChABC (0.5 U/mL, 0.5 mL per sample) for 12 h at 37 °C, the samples were washed three times with PBS. All samples were stained twice by Cy3-CHP (100 μM, 500 μL/sample) with shaking at 4 °C for 2 h. Subsequently, the samples were washed in PBS for 6 h and imaged with an iBright 1500 fluorescence imager. After the initial imaging, the samples were mechanically loaded according to their grouping. The loading speed was 10 mm/min; when the loading was maintained at 100 N for 10 s before decreasing to 0 N. The loading was cycled five times. These samples were stained again with Cy3-CHP (100 μM, 500 μL/sample) with shaking at 4 °C for 2 h and washed in PBS for 6 h. They were imaged again with the iBright 1500 fluorescence imager. The two imaging results were analyzed in parallel for ROI framing and quantification when the fluorescence increment (CHP fluorescence after loading - CHP fluorescence before loading) was determined for each sample.

**Relaxivity measurements**

T1 relaxivity of was measured by a 9.4 T small animal MRI scanner (Bruker BioSpec94/30 USR) with $^{1}$H planar receive-only surface coils (inner diameter: 20 mm, Bruker) at room temperature. Probes $Gd_n$-Cy5-CHP and $Gd_n$-Cy5-$^{S}$CHP were prepared into 200 μL solutions with $Gd^{3+}$ concentrations of 14, 28, 56, and 112 μM in water and scanned along with a serial dilution of Magnevist (gadopentetate dimeglumine). To obtain T1-weighted images of these sample solutions, T1 mapping sequences with the following parameters were employed: echo time (TE) = 7.0 ms; repetition time (TR) = 5500, 3000, 1500, 800, 400, and 200 ms; slice thickness: 0.5 mm; Field of View (FOV) = 30 × 25 mm; matrix dimensions = 128 × 128; bandwidth = ± 610.4 kHz; and echo train length (ETL) = 2; scanning time was 12 min 9 s. Plots of 1/relaxation time (1/T1, $s^{-1}$) versus $Gd^{3+}$ or peptide concentrations were produced to obtain their slopes as the relaxivity ($r_1$) values.

**Magnetic resonance (MR) imaging**

***MR imaging of CHP-stained, heat-denatured porcine osteochondral plug specimens***

Whole-depth articular cartilage from an adult pig's knee joints was fixed in 4% PFA for 48 h. The specimens were then rinsed with PBS before being dissected into small discs 6 mm in diameter with an osteotomy drill. Samples were rinsed with PBS during drilling for cooling. To denature the collagen content, the cartilage plugs were heated in a 100 °C water bath for 2 h. The heat-denatured specimens were placed in two EP tubes and incubated with a solution of $Gd_n$-Cy5-CHP or $Gd_n$-Cy5-$^{S}$CHP (5 μM, 5000 μL). Following a 96-hour incubation at 4 °C, the samples were washed three times for 3 h each with PBS to remove any unbound peptide The pieces were then fluorescently imaged using an IVIS Spectrum imager (PerkinElmer Lumina III) and scanned on a 9.4 T small animal MRI scanner (Bruker BioSpec94/30 USR) with a $^{1}$H planar receive-only surface coils (inner diameters 20 mm, Bruker) using a T1-mapping rapid acquisition with relaxation enhancement (T1-RARE) sequence. Heated and unstained cartilage plugs incubated in PBS were used as a blank control. Main imaging parameters: T1-RARE: echo time (TE) = 6.0 ms, repetition time (TR) = 200 ms, slice thickness: 0.7 mm, Field of View (FOV) = 20 × 10 mm, matrix dimensions = 192 × 96, bandwidth = 1400 Hz, scanning time was 48 s.

***In vivo MR imaging***

SD rats were imaged on a 9.4T small animal MRI scanner (Bruker BioSpec94/30 USR) with a $^{1}$H planar receive-only surface coil (inner diameters 20 mm, Bruker). Rats were anesthetized with 2% isoflurane in oxygen to maintain a constant respiration rate, kept warm by a thermal pad, and monitored by a small animal physiology-monitoring system. At the time of imaging, pre-injection baseline images as well as post-injection images were acquired for comparison. Probes ($Gd_n$-Cy5-CHP or $Gd_n$-Cy5-$^{S}$CHP) were injected intra-articularly into the knees at a dose of 1 nmol Gd/g body weight. Images were acquired at 6 h post-injection with Turbo rapid acquisition with relaxation enhancement sequence (TurboRARE, TSE). Main imaging parameters: T1WI (sequence: T1WI-3D-flash): echo time (TE) = 3.5 ms, repetition time (TR) = 16 ms, slice thickness: 0.2 mm, Field of View (FOV) = 22 × 22 × 8 mm, matrix dimensions = 192 × 192 × 40, scanning time was 18 min 50 s. T2WI (sequence: T2WI-2D-RARE): echo time (TE) = 27 ms, repetition time (TR) = 4000 ms, slice thickness: 0.2 mm, Field of View (FOV) = 25 × 25 mm, matrix dimensions = 228 × 228, scanning time was 7 min 36 s. The MR images were processed and analyzed by the RadiAnt DICOM Viewer software. The regions of interest (ROI) of the cartilage were drawn on eight sagittal slices to quantify signal intensity (SI). Additional ROIs were placed over the bone in the tibia, and an ROI was placed in the field of view without any tissue to measure the variation in background signal (air). The analysis was repeated for the pre-probe and post-probe MR data sets. To quantify signal enhancement in the cartilage, contrast to noise ratio (CNR) was calculated as:

$$CNR = (SI_{cartilage} - SI_{bone})/SD_{air}$$

Where SI is signal intensity, SD is the standard deviation. An average of all image slices was calculated for the pre-injection images ($CNR_{pre}$) and the post-injection images ($CNR_{post}$).[12]

**Clinical OA knee specimens**

All experimental plans involving knee specimens from OA patients were approved by the Ethics Committee of Nanjing Drum Tower Hospital (number: K228-1). These samples of femoral cartilage surgically removed from OA patients were fixed in 4% PFA solution for 48 h at room temperature and washed with PBS for 30 min three times. Osteochondral plugs with a diameter of 6 mm and a height of 5 mm were obtained from selected zooms of mild to severe cartilage degeneration using an osteotomy drill with continuous PBS rinsing for cooling. These plugs were incubated with $Gd_n$-Cy5-CHP (10 μM, 5 mL) at 4 °C for 2 days. After being washed with PBS 3 h three times, fluorescence and MR imaging were performed as described. After imaging, the $Gd_n$-Cy5-CHP labeled samples were decalcified in 20% EDTA for 7 days, dehydrated by 30% sucrose solution, and prepared for cryosections with a thickness of 10 μm. The sections were stained with safranin O and microscopically imaged.

A trapezoid-shaped piece of human OA femoral cartilage (short base × long base × height: 1 × 2.5 × 4 cm) was fixed in 4% PFA solution for 96 h and washed with PBS for 3 h three times. The sample was incubated with $Gd_n$-Cy5-CHP (10 μM, 40 mL) at 4 °C for 2 days. After being washed with PBS for 3 h three times, the sample was incubated with YO-PRO-1 working solution (Beyotime, C2025) for 2 days and washed with PBS for 1 h three times. Then the sample was decalcified in 20% EDTA (pH 7.0) solution for 2 weeks with the EDTA solution renewed every two days. The specimen was washed with PBS three times (30 min each round) before being decolorized with 25% Quadrol for 6 days. After being washed three times with PBS (1 h each round), the sample was delipidated with tert-butanol and dehydrated with 70% v/v tert-butanol, 27% v/v PEGMEMA500, and 3% w/v Quadrol for 6 days (with the solution changed every two days). Finally, the sample was immersed in a clearing medium (BB-PEG) made by 75% v/v benzyl benzoate (Sigma-Aldrich W213802), 22% v/v PEGMEMA500, and 3% w/v Quadrol for 4 days (with the solution changed every two days). The cleared sample was scanned in an imaging chamber filled with the BB-PEG medium using a LaVision Biotec Ultramicroscope II with a magnification of 1 (12 for local magnification) following the above-mentioned light-sheet microscopy procedures.

**Data and statistical analysis**

The data were analyzed and plotted using the GraphPad Prism 9 software. Numbers were displayed as the mean ± standard error of the mean (s.e.m.; normally distributed data) or median with interquartile range (non-normally distributed data, i.e., figure 3F) as indicated in figure legends. A paired *t*-test was used to compare two groups of paired normally distributed data. Unless otherwise noted in the figure legends, for multiple comparisons among group means of normally distributed data with homoscedasticity, one-way analysis of variance (ANOVA) with a *post hoc* Tukey HSD test was utilized. Datasets with non-normal distributions were analyzed using the Kruskal-Wallis test with a *post hoc* Dunn's test (figure 3F). *P* values below 0.05 were regarded as statistically significant, and *P* values above 0.05 were not shown in any figure. The Pearson correlation coefficient (r) was used to evaluate the correlation and test the null hypothesis ($H_0$), which states that there is no true correlation between the groups; the data sets appear to be more strongly correlated when the r value is closer to 1 and the *P* value is smaller. Statistical methods, *P* values, and sample size are indicated in figures and figure legends.

### Supplementary Table

**Table S1.** Fluorescently labeled peptides used in this study and their MALDI ms information.

| Peptide | Sequence | m/z calculated | | m/z found |
|---|---|---|---|---|
| Cy3-CHP | Cy3-Ahx-GPOGPOGPOGPOGPOGPOGPOGPO | [M + H]$^+$ | 3133.4 | 3134.0 |
| Cy3-$^{S}$CHP | Cy3-Ahx-PGOGPGPOPOGOGOPPGOOPGGOOPPG | [M + H]$^+$ | 3133.3 | 3132.4 |
| Cy5-CHP | Cy5-Ahx-GfOGfOGfOGfOGfOGfOGfOGfO | [M + Na]$^+$ | 3343.3 | 3343.8 |
| Cy5-$^{S}$CHP | Cy5-Ahx-OfGGOfGfGfOfOGOfGOOfGGOOffG | [M + Na]$^+$ | 3343.3 | 3343.3 |

Ahx: aminohexanoic acid, O: (2S, 4R)-4-hydroxyproline, f: (2S,4S)-4-fluoro-proline, Cy3: sulfo-Cyanine 3, Cy5: sulfo-Cyanine 5

### Supplementary Figures

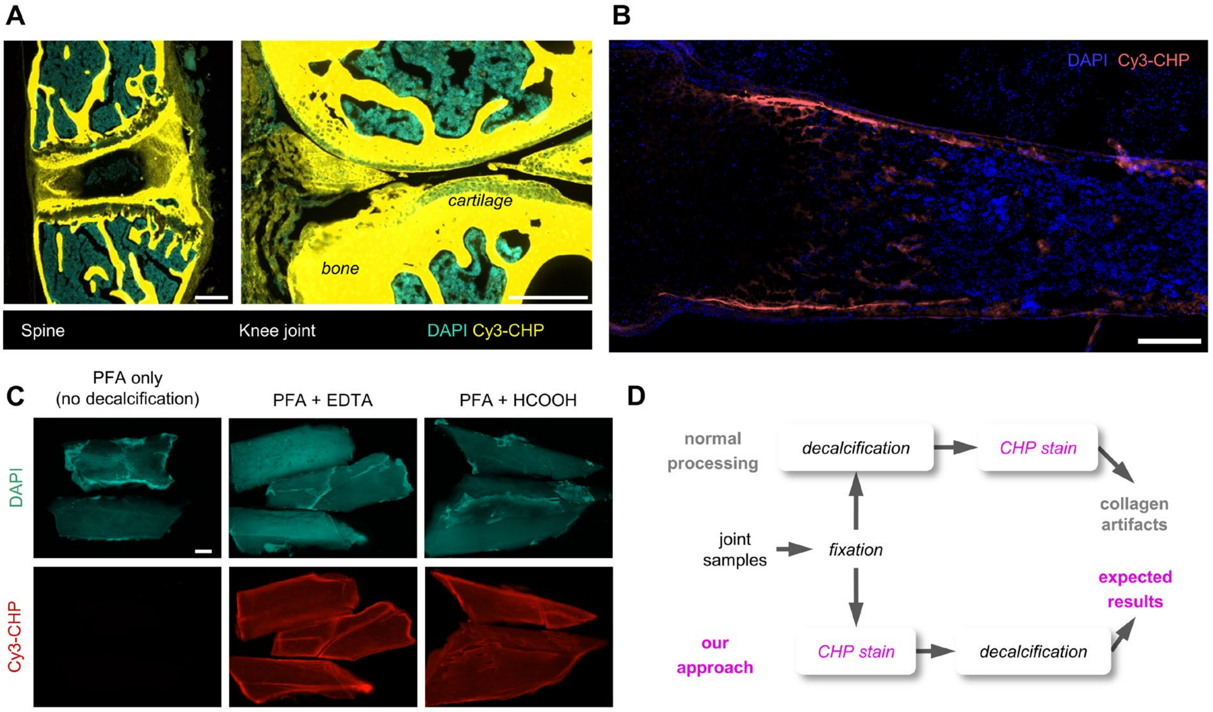


**Figure. S1. Modifications to the decalcification process of musculoskeletal tissues for CHP staining.** (**A**) Fluorescence micrographs of the spine and knee joint sections from normal mice stained with Cy3-CHP (15 μM in PBS, overnight) showed unexpected overwhelming signals in bone. These musculoskeletal specimens underwent a standard decalcification process, whereby the samples were fixed by 4% paraformaldehyde (PFA) and decalcified in EDTA before being cryosectioned in OCT compound embedding. **(B)** A representative fluorescence micrograph of a cryosection of the femur from a mouse at post-natal day 1 (P1) stained with Cy3-CHP (15 μM in PBS, overnight at 4 °C) showed signals only in the developing zones of endochondral ossification, where the collagen matrix undergoes protease-mediated remodeling and denaturation.[13] Because the femur was not fully calcified at P1, it could be directly cryosectioned without decalcification. The drastic difference in CHP fluorescence levels between (**A**) and (**B**) triggered us to investigate whether the decalcification process affects CHP binding to the bone. **(C)** Pieces of bone specimens from normal rat femurs pre-fixed with 4% PFA for 24 h were decalcified with EDTA (10% w/v in water, 24 h) or formic acid (10% v/v in water, 24 h) before stained with Cy3-CHP (5 μM in PBS, 24 h). The strong CHP fluorescence from the decalcified samples shows that the EDTA and formic acid decalcification treatments can both drastically promote CHP binding to the bone matrix and produce an artifact. We speculate that the decalcification process may cause the collagen molecules in the bone to denature. **(D)** To prevent this artifact in the histology of musculoskeletal samples, we modified the common specimen processing by performing CHP staining before the decalcification step. This modified approach has enabled us to preserve and interrogate the native molecular structure of collagen in the joint samples histologically. See Fig. 1A for representative results from the two different procedures and Supplementary Materials and Methods for details. Scale bars: 300 μm (**A**), 200 μm (**B**, **C**).

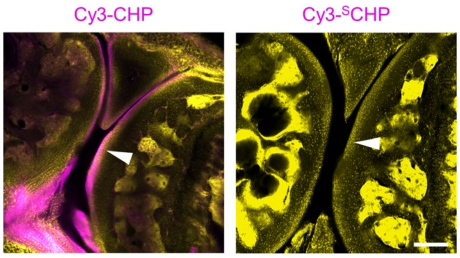


**Figure. S2. Collagen hybridization in joint tissues relies on the triple-helical folding capacity of CHP.** Cryosections of the normal C57BL/6 mice's knee joints showed clear fluorescence signals (magenta) in the cartilage (arrowheads) and meniscus when stained with Cy3-CHP but not the sequence-scrambled control peptide (Cy3-$^{S}$CHP). Yellow: DAPI. Scale bar: 200 μm.

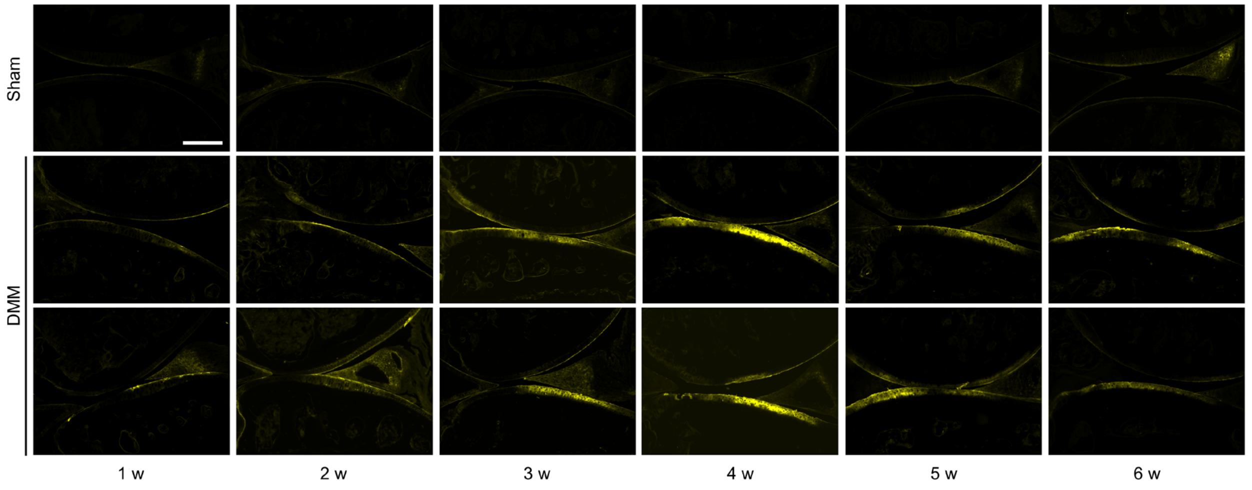


**Figure. S3. Additional fluorescence images of the mice's post-operation joint sections.** Micrographs of the knee cartilage cryosections prepared from the mice (*n* = 3) at each time point post-DMM- or sham-operation stained with Cy3-CHP. The quantification of the fluorescence intensity from each CHP-stained slide and its corresponding OARSI grade are plotted in Fig. 1D. Scale bar: 300 μm.

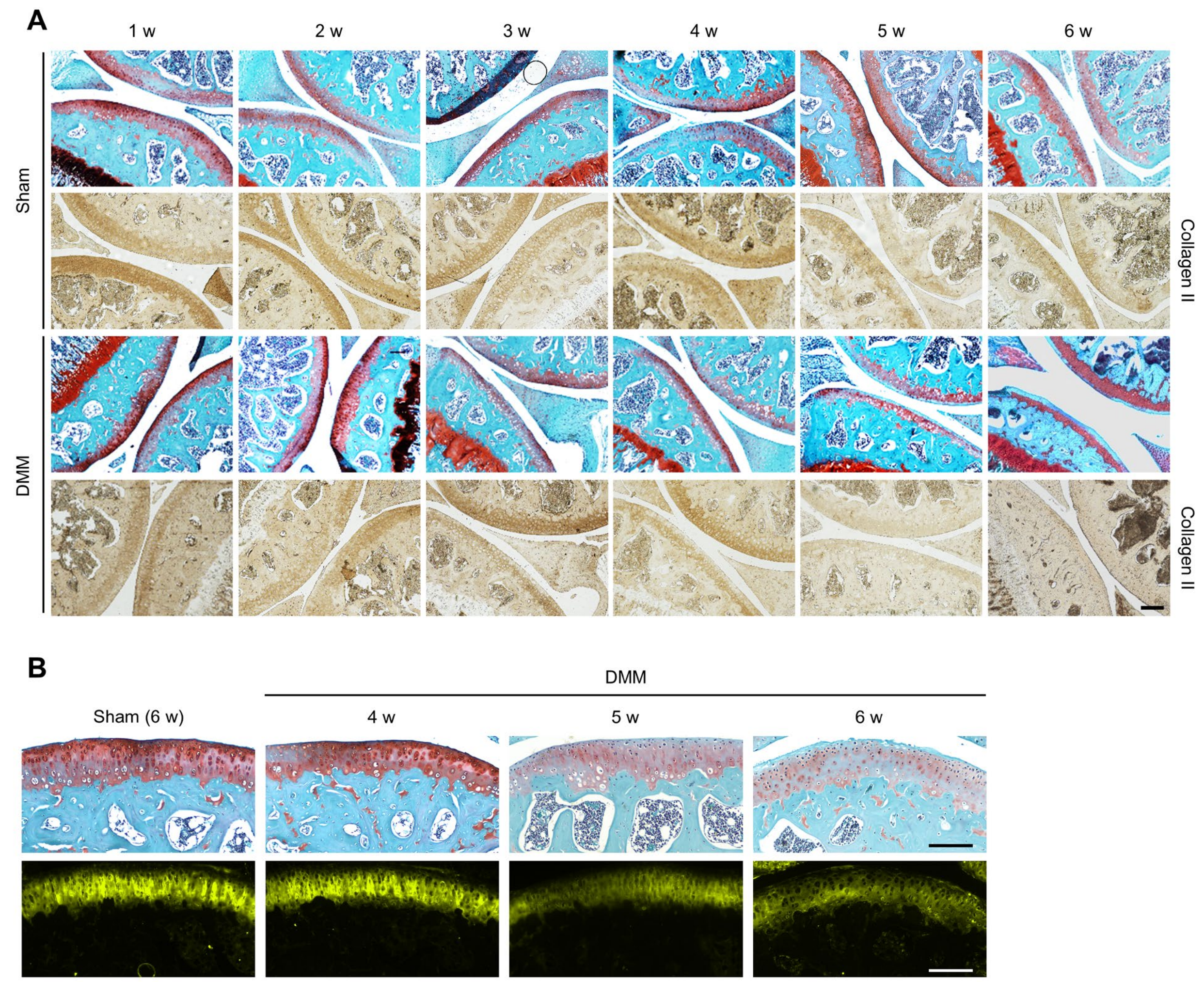


**Figure. S4. Loss of collagen II in the mice's DMM-injured knee cartilage.** (**A**), (**B**) Representative images of knee joint paraffin-embedded sections prepared from the DMM-injured mice at each time point post-operation, stained with Safranin-O/fast green (top row) or anti-collagen II antibodies (detected by HRP-immunohistochemistry, bottom rows in **A**, or by a fluorescently labeled secondary antibody, bottom row in **B**). Once the OARSI grade exceeds 4 (i.e., in weeks 5 and 6 post-DMM), the cartilage's collagen II content is massively reduced. This trend strongly correlates with the diminishing CHP fluorescence intensity and the destructed morphology (Fig. 1B). Scale bar: 300 μm (**A**), 138 μm (**B**).

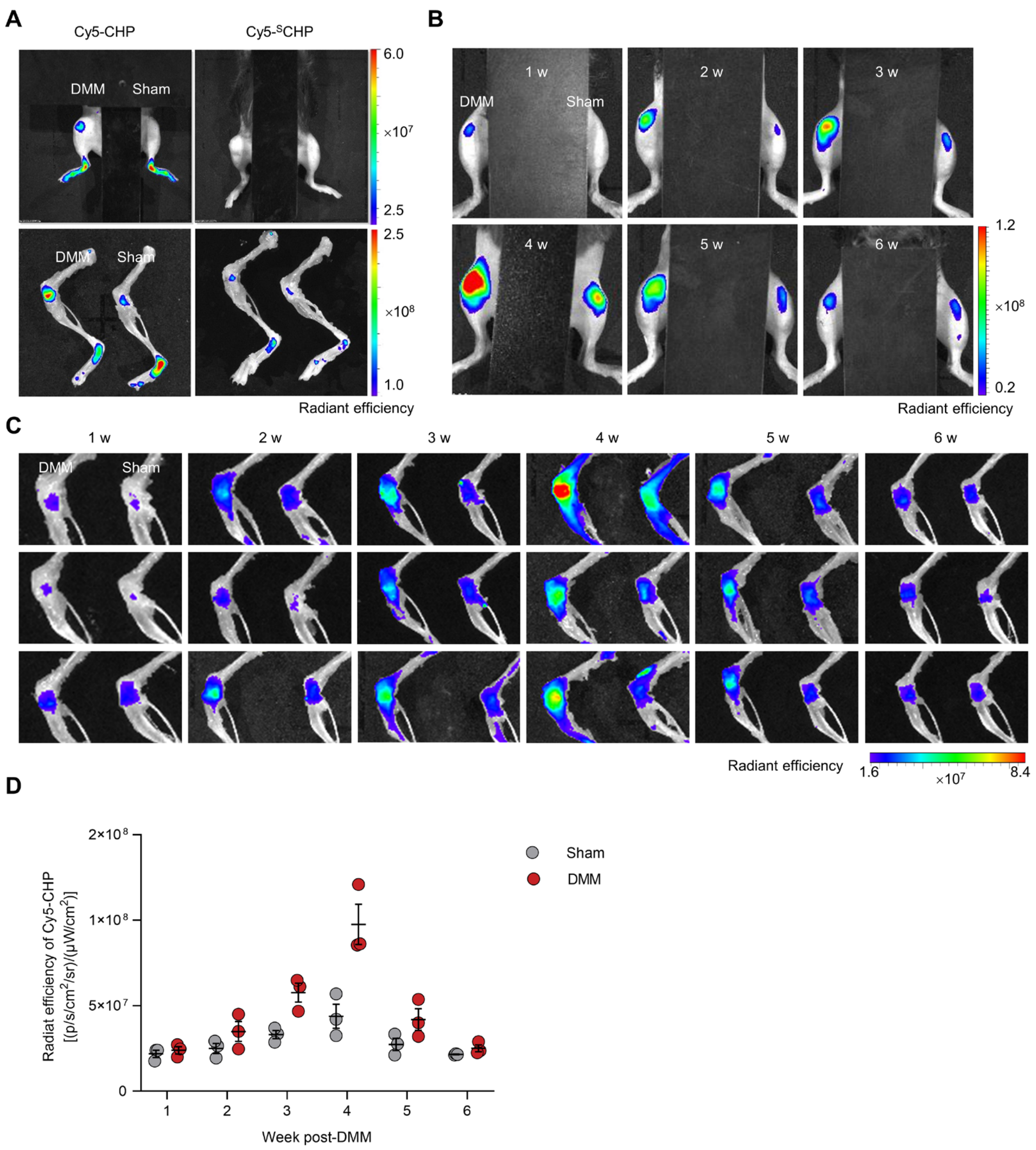


**Figure. S5. In vivo CHP targeting of collagen denaturation in mouse knees during OA progression.** (A) Representative fluorescence images of living mice and their skeletal tissues 4 weeks post-operation showed a robust fluorescence signal in the DMM-injured knee compared to the sham-operated knee for Cy5-CHP, but not the sequence-scrambled control peptide Cy5-$^{S}$CHP. (**B**) Representative live fluorescence images of the mice at all time points post-operation. The CHP fluorescence intensity from the DMM-operated knee was consistently higher than the sham control of the same mouse at all times. (**C**) Ex vivo near-infrared fluorescence images of the isolated knee joints from all the mice at all time points post-DMM [$n$ = 3 mice per week; DMM (left) versus sham (right) from the same mouse are shown in each image]. All images in this figure (**A** to **C**) were acquired 3 h post intravenous injection of 1 nmol of Cy5-CHP or Cy5-$^{S}$CHP with the probe fluorescence shown in rainbow color as radiant efficiency [unit: (p/sec/cm$^2$/sr)/(μW/cm$^2$)]. (**D**) Total Cy5-CHP fluorescence intensities quantified from all isolated knee joints in **C**.

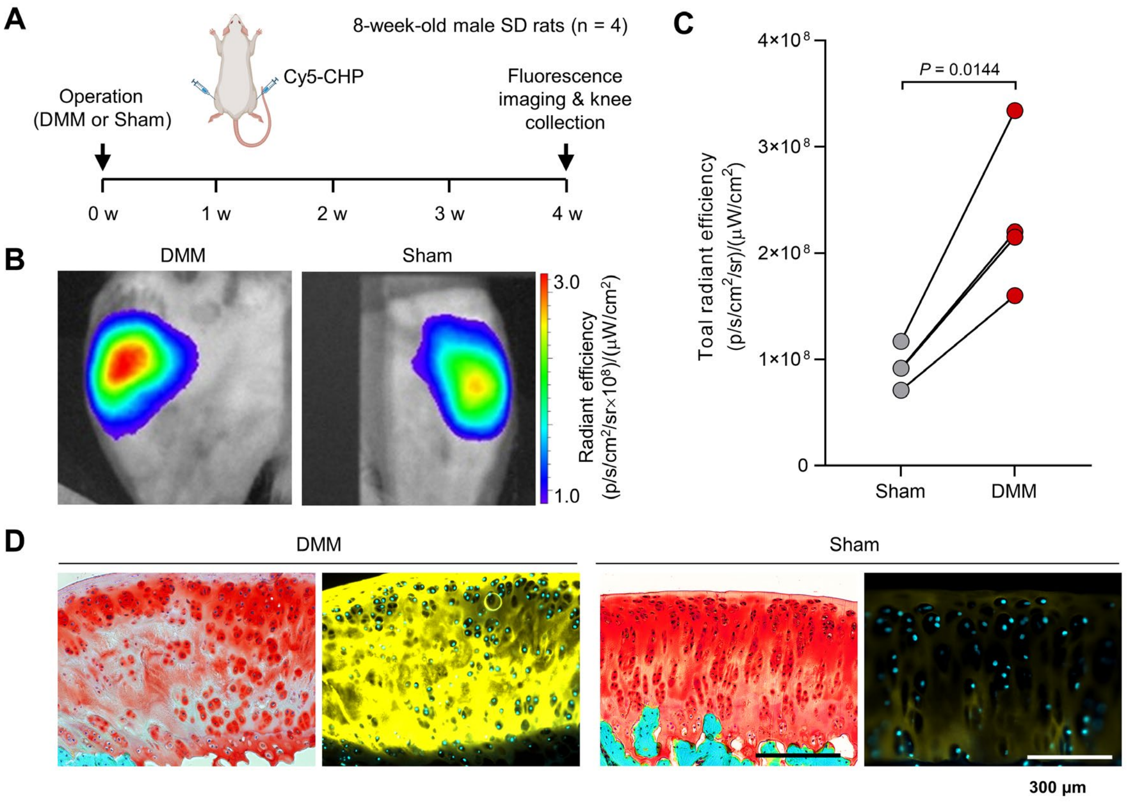


**Figure. S6. Cy5-CHP reveals collagen denaturation in knee cartilage of osteoarthritic rats in vivo.** (**A**) Timeline of the experiment on Sprague Dawley (SD) rats following intra-articular injection of Cy5-CHP. The OA model was prepared by DMM surgery in the right knees of these rats with a sham-operation in their left knees. (**B**) Representative in vivo images of the Cy5-CHP fluorescence from the DMM- or sham-operated knee areas of the SD rats 4 weeks post-operation. The images were obtained 3 h post-CHP injection. (**C**) Quantified Cy5-CHP fluorescence signals from the paired knee joints isolated from the rats in (**B**). Data were statistically analyzed using a paired t-test. (**D**) Representative fluorescence images of the cryosections of the knee cartilage collected from (**B**) showing in vivo uptake of Cy5-CHP specifically binding to the DMM-injured cartilage. For comparison, a neighboring section of each tissue sample was stained with Safranin-O/fast green, which showed mild GAG loss without visible erosion (OARSI grade < 3) in the cartilage of the DMM-operated knee. These images demonstrated that Cy5-CHP can identify molecular collagen denaturation in the rat knee cartilage without apparent morphologic alteration during early-stage OA. Also, in the DMM-injured cartilage, the areas of high Cy5-CHP fluorescence match closely with the regions showing reduced GAG contents in the Safranin O stain. Scale bars: 300 µm (**D**).

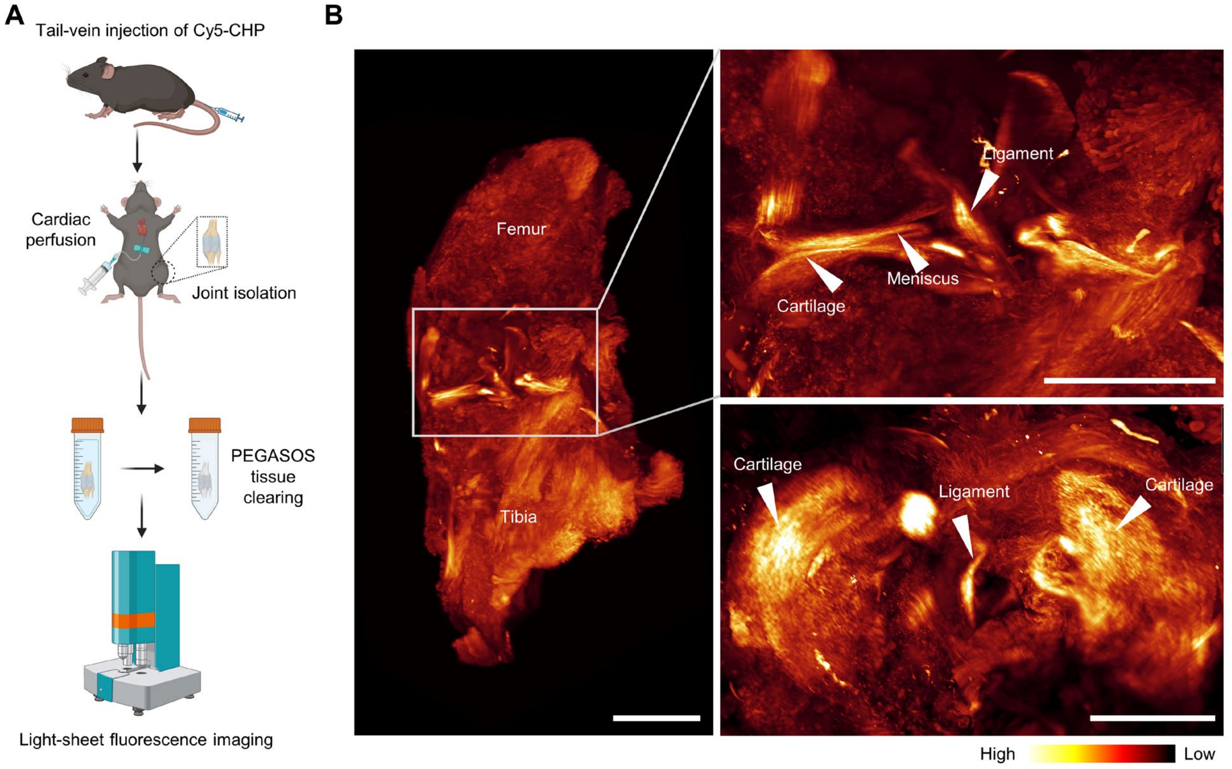


**Figure. S7. Tissue clearing of mouse knee joints for light-sheet fluorescence microscopy**. (**A**) Tissue clearing process of a knee joint specimen collected from a mouse pre-injected with Cy5-CHP in vivo (detailed in Supplementary Materials and Methods). (**B**) Light-sheet microscopy fluorescence images of a normal knee joint harvested from a healthy mouse 3 h post intravenous injection of 1 nmol of Cy5-CHP. The fluorescence images showed detectable but weak CHP binding in the normal cartilage, ligaments, and meniscus, indicating a low physiological level of molecular collagen denaturation in normal mouse joints. The low fluorescence signals were digitally enhanced in these images to allow visualization. These images are representative of similar results from 3 mice. Scale bars: 1 mm.

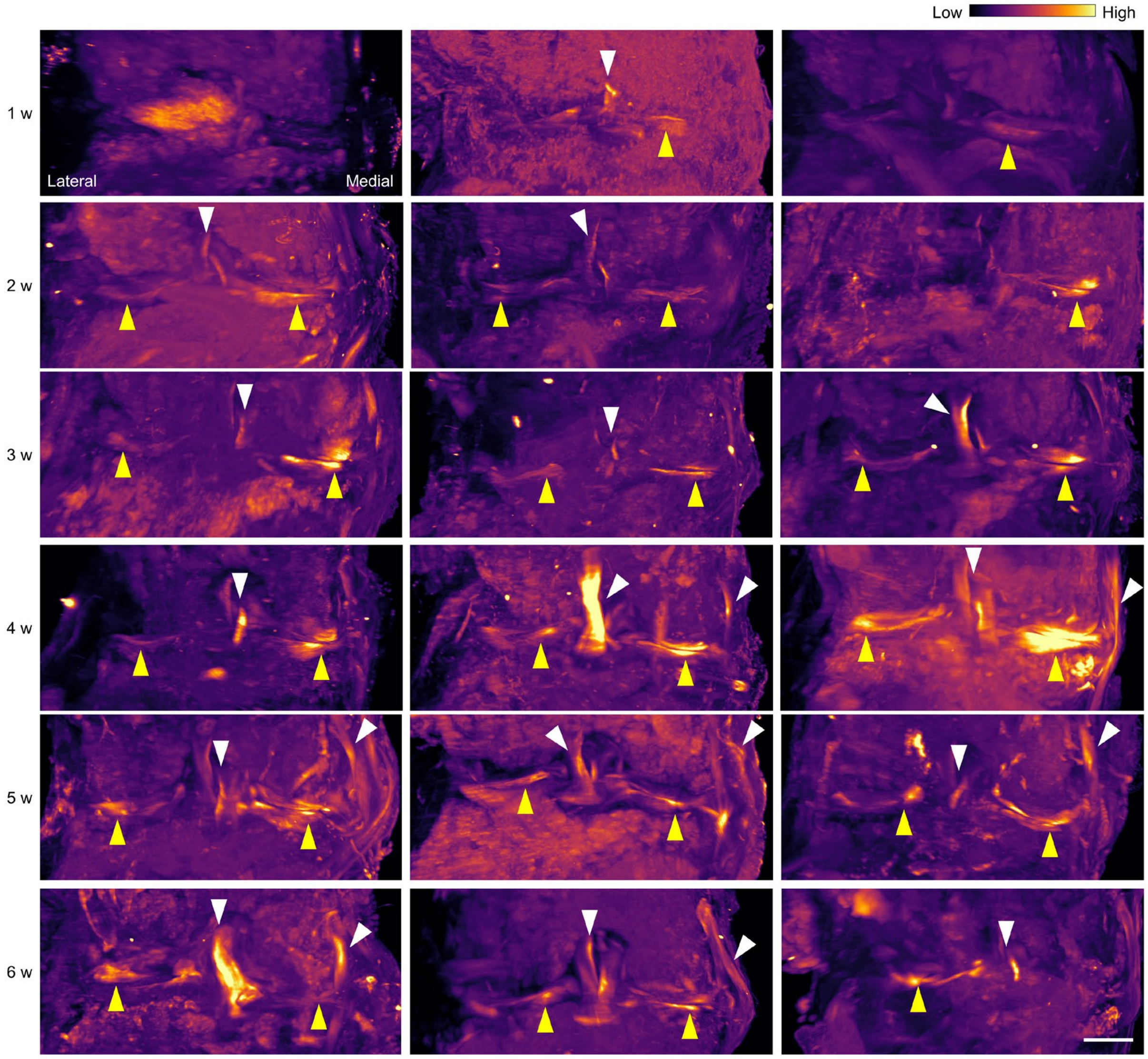


**Figure. S8. Compressed three-dimensional light-sheet fluorescence microscopy images of the mouse knee joints from 1 to 6 weeks post-DMM-surgery.** The joint specimens were harvested and cleared 3 h post-intravenous injection of Cy5-CHP ($n$ = 3 mice per time point). Yellow arrowheads: cartilage, white arrowheads: ligaments. Scale bar: 500 μm.

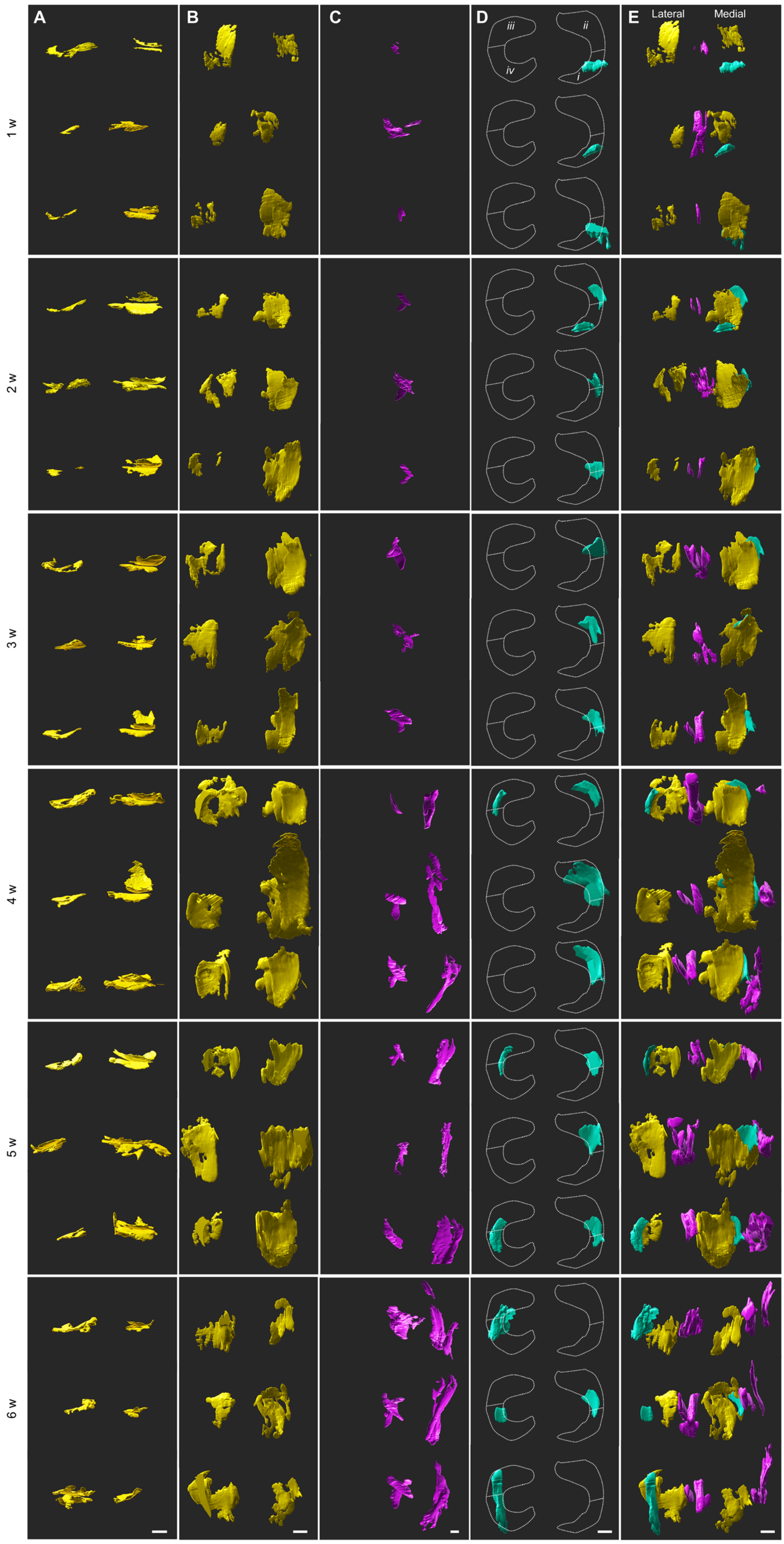


**Fig. S9. Three-dimensional reconstruction of the mice's diseased knee joints at all time points post-DMM-operation showing the lesion volumes with molecular collagen defects in each tissue.** Higher-than-background Cy5-CHP fluorescence signals were picked and color-labeled according to the tissue type in individual displays (**A** to **D**) and overlaid images (**E**): cartilage (yellow, **A** and **B**), ligaments (magenta, **C**), and meniscus (cyan, **D**). Cy5-CHP was intravenously injected into the mice ($n = 3$ mice per time point) 3 h before the joints were collected and cleared for light-sheet fluorescence microscopy. Frontal view (**A**), top view (**B** to **E**). Scale bars: 300 µm.

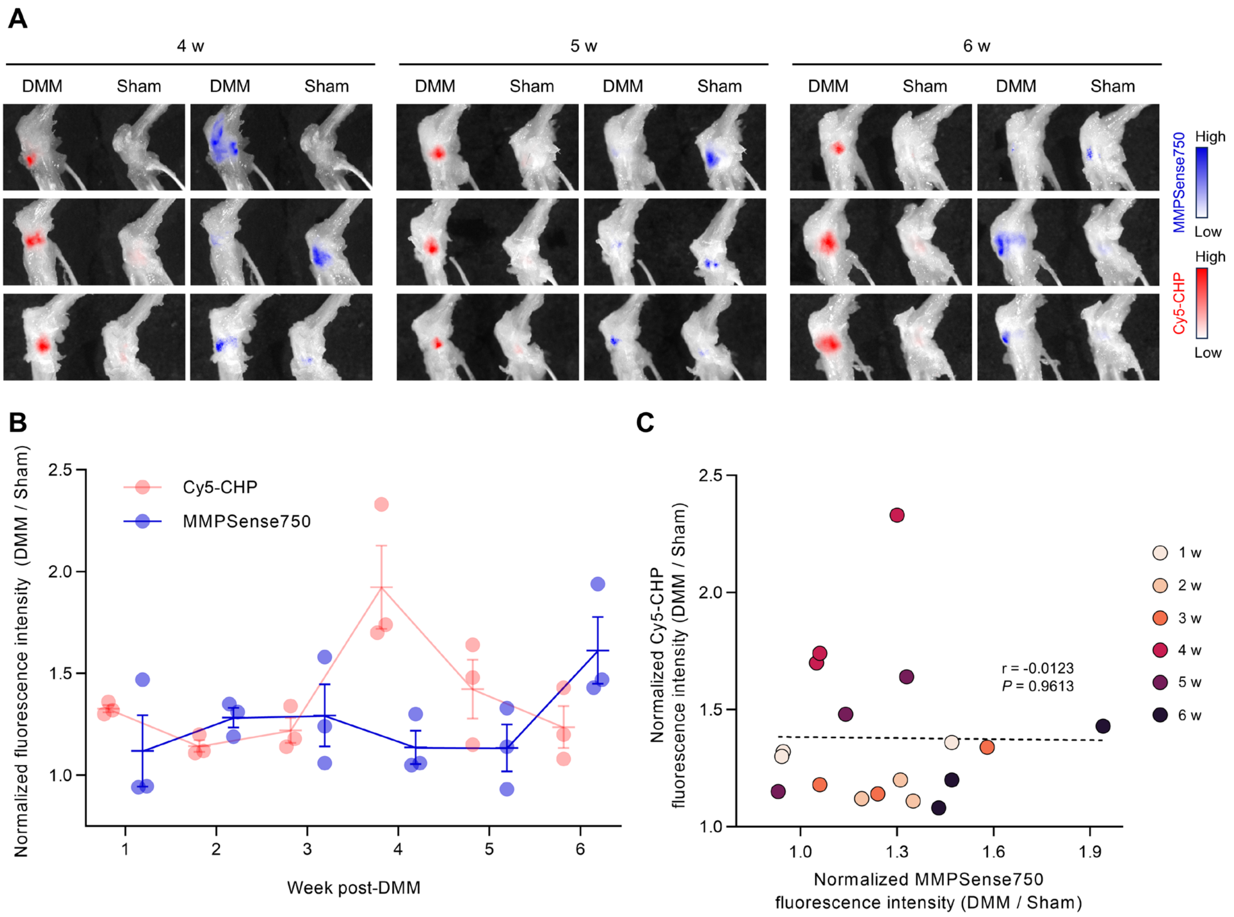


**Figure. S10. In vivo fluorescence profiles of MMPSense and Cy5-CHP.** (**A**) Up-close ex vivo fluorescence images of all mice's knee joint pairs, harvested 4-, 5-, and 6-week post-operation, highlighting highly distinct locations of in vivo fluorescence signals from the intravenously injected Cy5-CHP (3 h p.i.) and MMPSense750 (24 h p.i.). Each image contains two knee joints from the same mouse, and each knee pair has been imaged and displayed twice (side by side, left image: Cy5-CHP, right image: MMPSense750). The fluorescence intensity display of each image has been adjusted such that a minimal signal is shown in either of the knees with a lower fluorescence level. (**B**) Normalized fluorescence intensity of MMPSense750 and Cy5-CHP of each mouse's DMM-injured knee (i.e., fold change over its sham-operated counterpart) at all time points. A ratio below or equal to 1.0 indicates that the MMP activity or CHP uptake in the DMM-operated knee is no higher than the sham. Data are mean ± s.e.m. (**C**) The normalized fluorescence intensities of Cy5-CHP and MMPSense750 showed a poor correlation between the two data sets according to the Pearson correlation coefficient (r).

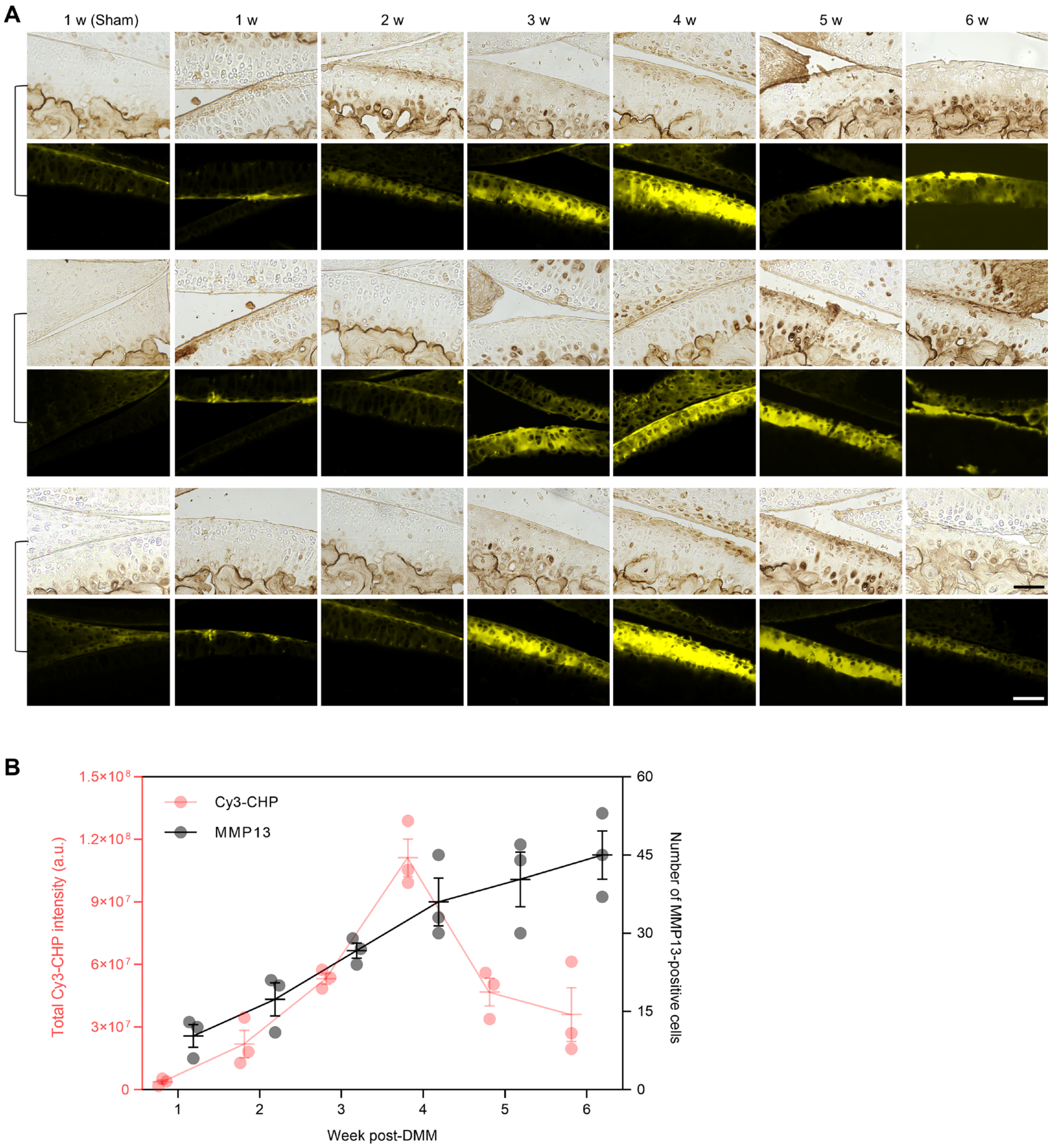


**Figure. S11. Double staining of denatured collagen (Cy3-CHP) and MMP13.** (**A**) Paired micrographs of sections of all mice's DMM-operated knee joints, harvested weekly post-operation, double stained with an anti-MMP13 antibody (immunohistochemistry) and Cy3-CHP (fluorescence). Each pair of images represents one mouse knee. (**B**) Numbers of MMP-positive cells and the Cy3-CHP fluorescence intensities in the joint sections (counted or quantified from **A**), plotted against the time post-DMM operation. These histologic images and quantification showed distinct spatiotemporal profiles between MMP13 expression and collagen denaturation in the DMM-induced OA cartilage. Data are mean ± s.e.m. (**B**). Scale bars: 70 µm (**A**).

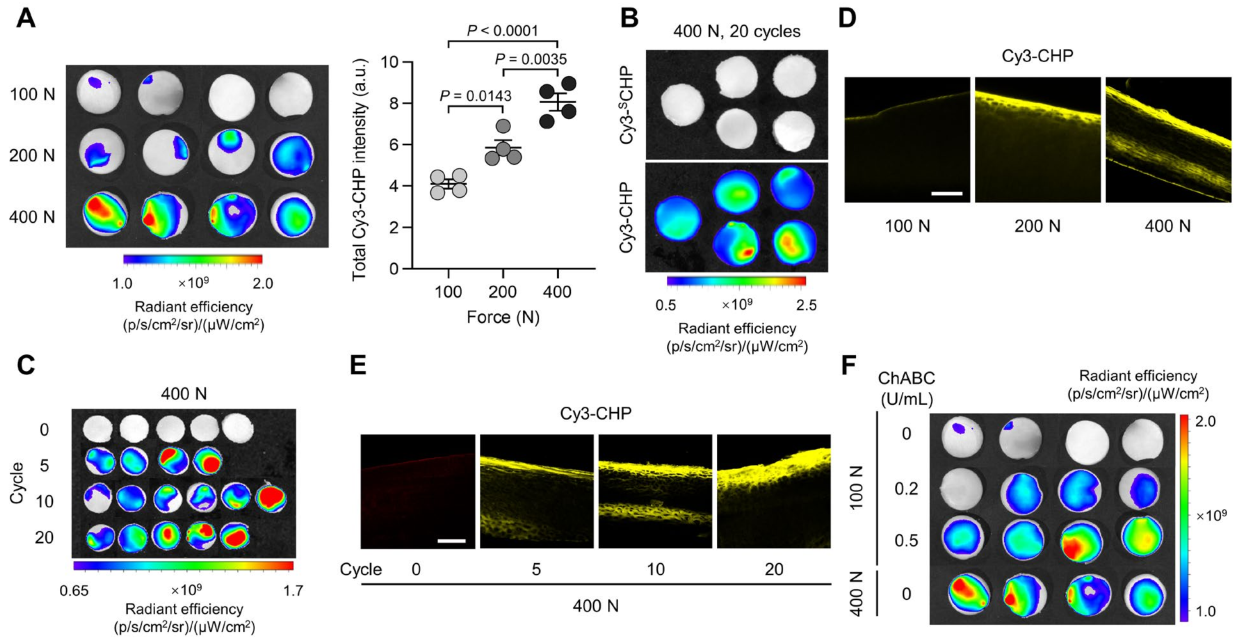


**Figure. S12. Supplementary mechanical testing results.** (**A**) Fluorescence images (left) and quantification (right) of groups of cylinder-shaped normal porcine osteochondral plug specimens (diameter: 1 cm, $n = 4$ samples) stained with Cy3-CHP after 10 cycles of compressive loading of 100, 200, or 400 N (i.e., 1.3, 2.5, or 5.1 MPa) on the cartilage. Data are mean ± s.e.m. and analyzed using one-way ANOVA followed by Tukey's multiple-comparison tests. (**B**) Fluorescence images of normal porcine plug specimens ($n = 5$ samples) subjected to 20 cycles of 400 N loading, stained with Cy3-CHP or Cy3-$^{S}$CHP. Denatured collagen was undetectable by the control probe Cy3-$^{S}$CHP with a scrambled peptide sequence (Cy3-Ahx-PGOGPGPOPOGOGOPPGOOGPGOOPPG), indicating that the CHP staining is specifically based on the triple-helix hybridization. (**C**) Fluorescence images of all Cy3-CHP-stained samples with 0-20 cycles of 400 N loading. Representative images and quantification are shown in Fig. 3F. (**D** and **E**) Confocal fluorescence microscopic scans of the cross-sections of the loaded cartilage samples from (**A**) (displayed in **D**) and (**C**) (displayed in **E**), showing strong Cy3-CHP staining close to the cartilage surface. Each image was representative of similar results from 4-6 samples within each group. (**F**) Fluorescence images of all the normal or ChABC-treated cartilage samples with 100 or 400 N loading stained with Cy3-CHP. Quantification is shown in Fig. 3G. Scale bars: 200 μm (**D** and **E**).

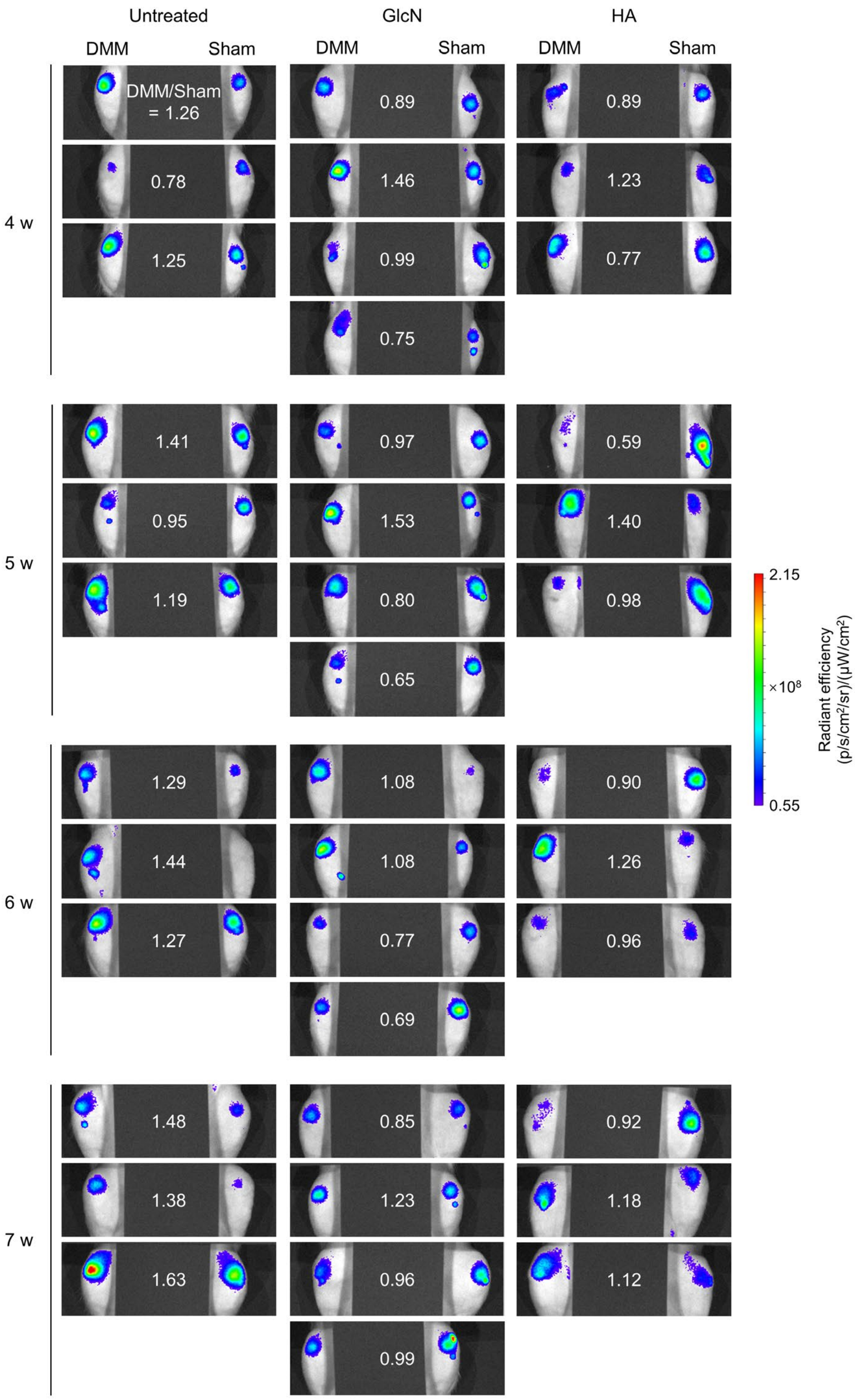


**Figure. S13. Fluorescence images of all rats from the untreated, GlcN-, and HA-treated groups 4–7 weeks post-DMM-operation.** The images were acquired 3 h post-intra-articular Cy5-CHP injection. The white number in each image shows the ratio of the quantified Cy5-CHP fluorescence signals between the DMM- and sham-operated knee joint of each rat.

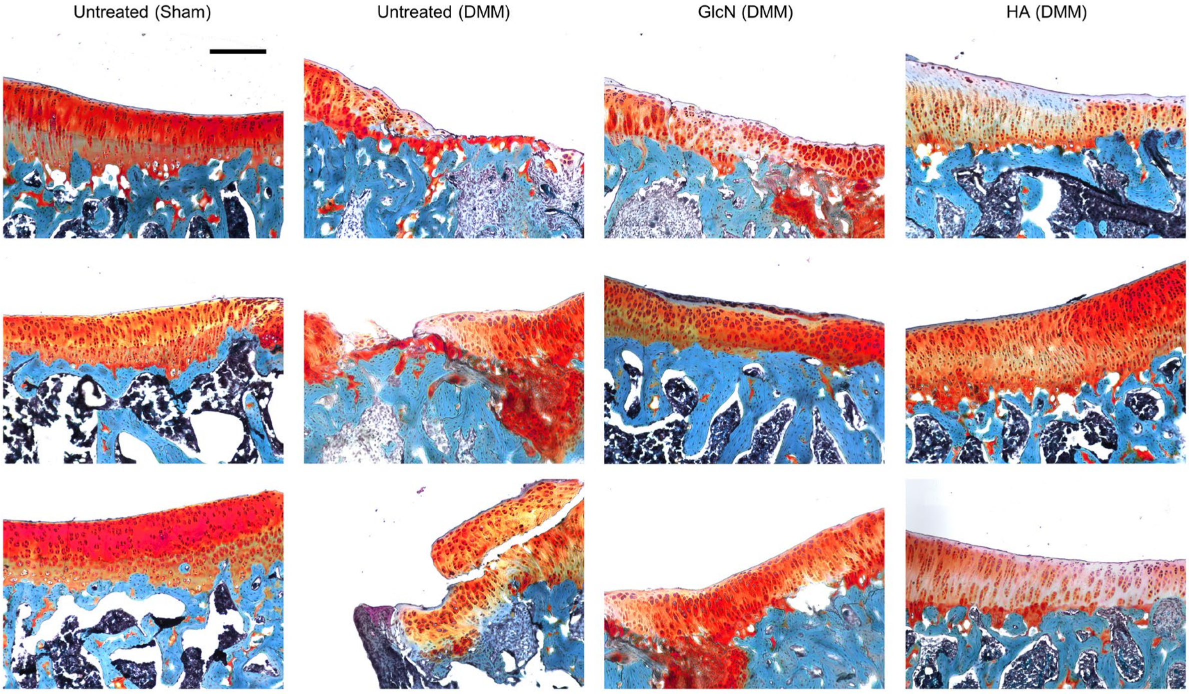


**Figure. S14. All micrographs of endpoint histology (Safranin O stain) at 7 weeks post-operation with or without treatments.** These images showed the integrity of the tibial plateau cartilage from the rats in the untreated, GlcN, and HA groups. Each image represents one rat in each group. Scale bar: 250 μm.

A

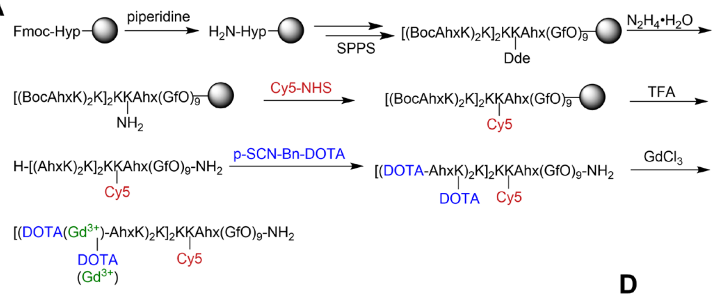


B

| | Number of $Gd^{3+}$ per molecule |
|---|---|
| $Gd_n$-Cy5-CHP | 5.33 ± 0.33 |
| $Gd_n$-Cy5-$^S$CHP | 6.33 ± 0.33 |

C

| | $r_1$ ($mM^{-1}$ $s^{-1}$) per molecule | $r_1$ ($mM^{-1}$ $s^{-1}$) per Gd ions |
|---|---|---|
| Gd-DTPA | 4.17 | 4.17 |
| $Gd_n$-Cy5-CHP | 35.22 | 6.60 |
| $Gd_n$-Cy5-$^S$CHP | 37.79 | 5.96 |

D

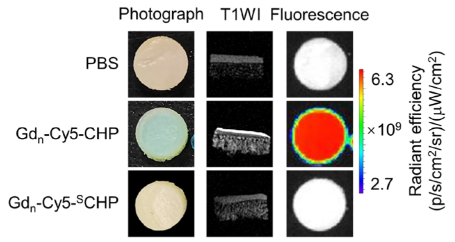


**Figure. S15. Synthesis and characterization of contrast agent $Gd_n$-Cy5-CHP.** (**A**) Synthesis route of $Gd_n$-Cy5-CHP. For the control agent $Gd_n$-Cy5-$^S$CHP lacking denatured collagen binding, the CHP peptide sequence was scrambled ($^S$CHP: OfGGOfGfGfOfOGOfGOOfGGOOffG). (**B**) The number of $Gd^{3+}$ ions labeled per peptide molecule calculated using $Gd^{3+}$ concentrations measured by inductively coupled plasma optical emission spectroscopy (ICP-OES). $n$ = 3 individual samples. Data are mean ± s.e.m. (**C**) T1 relaxivity of Magnevist (Gadopentetate Dimeglumine, Gd-DTPA), $Gd_n$-Cy5-CHP, and $Gd_n$-Cy5-$^S$CHP measured by 9.4 T small animal MRI scanner in vitro. (**D**) MR and fluorescence imaging of heat-denatured osteochondral plugs (diameter: 6 mm) excised from normal porcine patella-femoral joints treated with the $Gd_n$-Cy5-CHP probes. To denature the cartilage's collagen in the samples, paraformaldehyde-fixed porcine plug specimens were heated in boiling water for 2 h before being grouped into three to be incubated at 4 °C in 5 mL of PBS or $Gd_n$-Cy5-CHP/$Gd_n$-Cy5-$^S$CHP solutions ($n$ = 3 individual plugs per group, CHP/$^S$CHP probe concentration: 5 µM). Following extensive wash with PBS, the samples were fluorescently imaged with an IVIS spectrum imager and scanned by a 9.4 T MRI scanner. Under the bright field, the plugs incubated with $Gd_n$-Cy5-CHP were stained blue due to the Cy5 coloration. The fluorescence intensity of the porcine cartilage stained with $Gd_n$-Cy5-CHP was extremely high. Similarly, strong MR T1 enhancement was only visible from the group treated with $Gd_n$-Cy5-CHP in the sagittal cross-section of the samples scanned with a T1-RARE sequence. The lack of T1 and fluorescence signals in the specimens stained with $Gd_n$-Cy5-$^S$CHP indicated that triple-helix hybridization is specifically required for the contrast agent's binding to denatured collagen. Each image represents similar results from the three samples within each group.

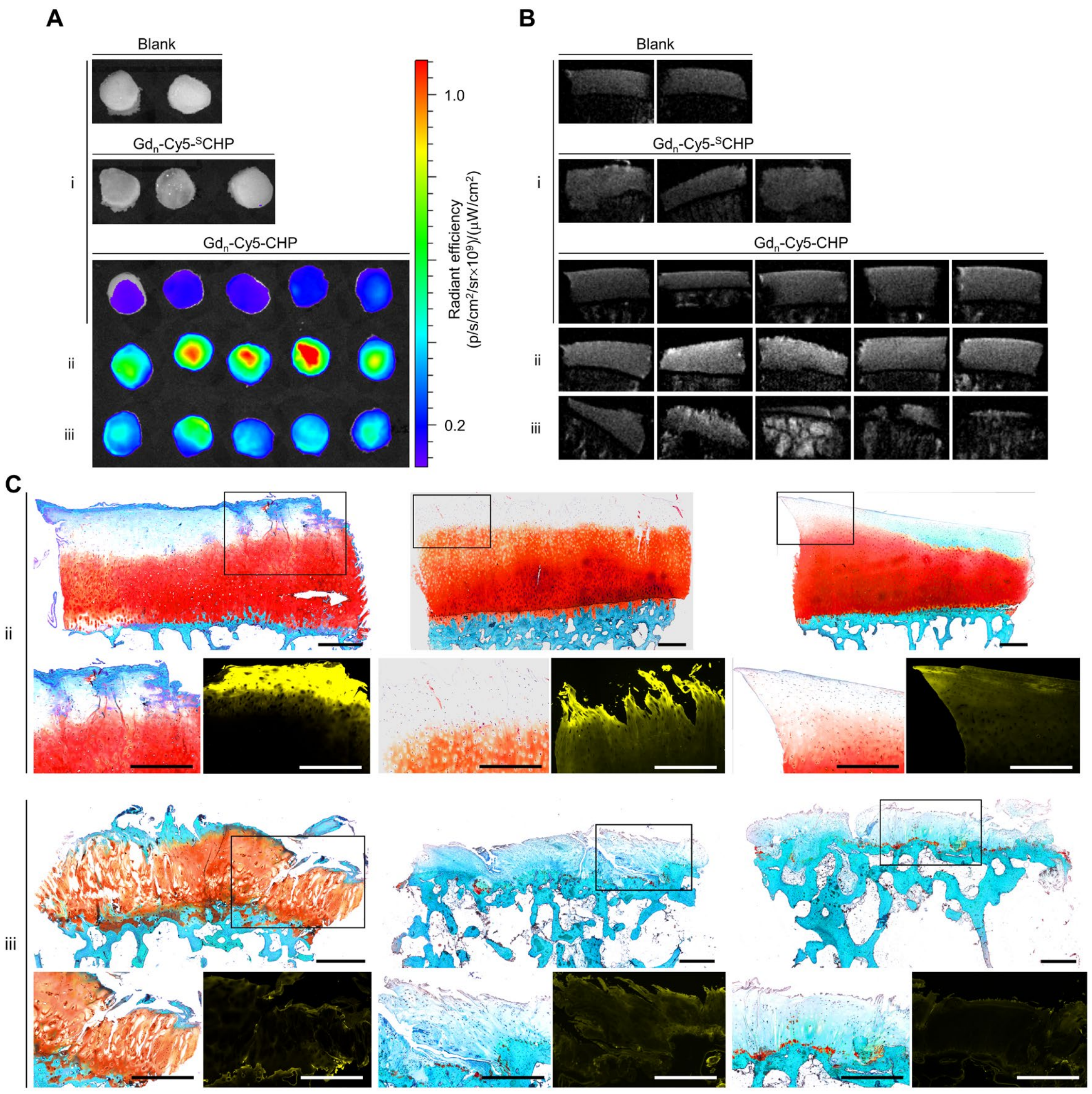


**Figure. S16. Ex vivo fluorescence (A) and MR T1WI imaging (B) of clinical OA osteochondral plug specimens (diameter: 6 mm) excised from regions with mild-to-severe cartilage degeneration (groups i-iii) labeled with Gd$_n$-Cy5-CHP or Gd$_n$-Cy5-$^S$CHP.** The samples treated with the non-collagen-hybridizing control probe Gd$_n$-Cy5-$^S$CHP were indistinguishable from the blank samples without detectable fluorescence and T1WI signal enhancement. This result highlighted that the CHP's contrast enhancement is specifically due to its triple-helical collagen hybridization. (**C**) Additional Safranin O stain of the cryosections of the clinical specimens with moderate-to-severe cartilage degeneration from (**A** and **B**). Each boxed area is displayed in magnification (left) along with in situ fluorescence from the tissue-bound Gd$_n$-Cy5-CHP (right) below each whole-section scan. Scale bars: 500 μm (**C**).

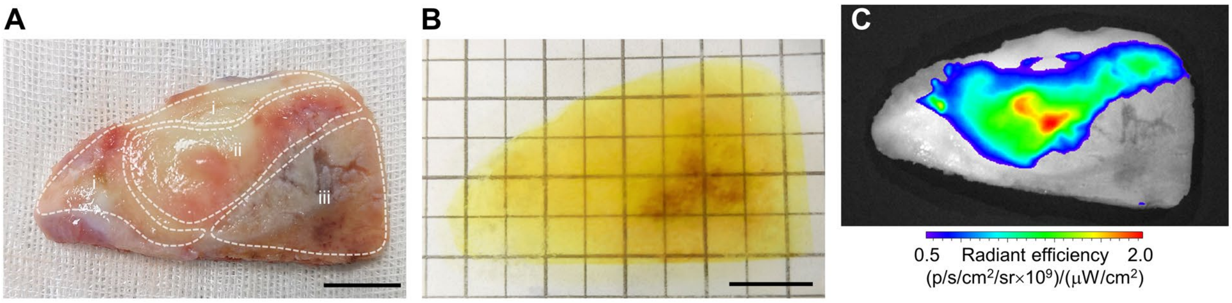


**Figure. S17. Photographs of femoral knee cartilage surgically removed from an OA patient before (A) and after Cy5-CHP staining and tissue clearing (B)**. i-iii: Regions with mild-to-severe levels of cartilage degeneration and OARSI grades (degeneration levels: i, mild; ii, moderate, iii, severe). (**C**) Bulk fluorescence image of the sample showing areas with strong Cy5-CHP binding. Light-sheet fluorescence imaging of this life-size specimen is in Fig. 6G. Scale bars: 1 cm (**A** and **B**).
